# A rRNA hybridization-based approach for rapid and accurate identification of diverse fungal pathogens

**DOI:** 10.64898/2026.03.06.26347616

**Authors:** Elizabeth A. Yee, Brooks J. Burt, Poppy C. S. Sephton-Clark, M. Lauren Donnelly-Morell, Aidan Z. Wang, Isaac H. Solomon, Evan Mojica, Christina A. Cuomo, Roby P. Bhattacharyya

## Abstract

Invasive fungal infections are a global threat for which early diagnosis is critical for patient outcomes, but current diagnostic measures remain notoriously slow. Here we extend our multiplexed, hybridization-based rRNA-targeted strategy for rapid, sensitive pathogen identification, previously designed for diverse bacteria and *Candida* species, to identify diverse fungal pathogens. We created a set of 91 probes targeting 86 medically relevant fungal species, designed to recognize regions of differential conservation across taxonomic groupings, from class-to species-specific probes. We assessed assay performance across a Training Set of 93 clinical isolates spanning 32 species of common fungal pathogens across 18 genera, with Pearson correlations of probeset reactivity profiles identifying the pathogen at the species, genus, and family level with 83%, 94%, and 95% accuracy, respectively, in a leave-one-out analysis. We developed a new classifier on this Training Set, using taxonomic categories to select progressively more informative probes at each taxonomic level. After optimization, we assessed performance on an independent Validation Set of 54 clinical isolates spanning the same species as the Training Set, with 91%, 94%, and 98% at the species, genus, and family levels, respectively. We piloted our assay on formalin-fixed paraffin-embedded (FFPE) tissue, demonstrating rapid, culture-independent fungal identification from this high-value clinical sample type, often the sole specimen available. The assay requires <30 minutes hands-on time (or <65 minutes from FFPE tissue), returning results in <8 hours from cultured specimen on an RNA detection platform available in clinical laboratories.

**Importance:** Timely identification of fungal pathogens is critical for patient outcomes. Here we describe a multiplexed hybridization-based assay targeting rRNA that capitalizes on the high sequence conservation and abundance of rRNA to generate unique reactivity profiles that serve as “fingerprints” of diverse fungal pathogens. By comparing these fingerprints to a Training Set of known isolates, this novel assay enables accurate identification of 32 species of pathogenic fungi from crude lysates of cultured clinical isolates.

## Introduction

Invasive fungal infections (IFIs) are a growing threat to human health, accounting for 1.6 million global deaths annually (1). Advances in immunosuppressive drugs and chemotherapy have resulted in larger immunocompromised populations vulnerable to opportunistic pathogens (2–4). Early diagnosis of fungal infections is critical for patient outcome, but is notoriously slow, relying heavily on expert clinical mycologists identifying morphology from cultured or fixed patient specimens (1, 5, 6). As such, healthcare systems are in need of rapid and accurate diagnostic tools to identify fungal infections (3–5).

Progress has been made in fungal diagnostics using several different modalities. Serological testing can help diagnose IFIs by detecting fungal antigens, or host antibodies reflecting exposure, but many assays have cross reactivity (7, 8). Serology platforms often have a quick turnaround time and no culturing requirement, but are limited by a narrow species range or high false positive rate for more general fungal biomarkers such as 1,3-β-D-glucan (3, 7, 9, 10). Widely used for bacterial identification, matrix assisted laser desorption/ionization time of flight (MALDI-TOF) mass spectrometry is gaining traction as a method of fungal identification (7, 11–13). Although this method is easy to run, it requires culture growth which is not always possible, and identification is complicated by variable surface proteome expression across the fungal life cycle, making fungi harder to identify against reference databases (5, 7, 14).

As costs fall and efficiency increases, sequencing is increasingly used for fungal species identification. Targeted amplicon sequencing of the internal transcribed spacer region (ITS), ribosomal RNA (rRNA) genes, or additional highly conserved genes has been widely adopted by reference laboratories to discriminate between fungal species (5, 9, 15, 16). However, fungal DNA in primary clinical samples is often very scarce, requiring amplification that can result in false positives from low-level contaminants (17). Conversely, false negatives can occur due to inhibition of PCR enzymes (17). Despite these improvements in fungal diagnostics, no single strategy or commercial test can identify a wide variety of fungal organisms rapidly and accurately. Additionally, sequencing assays are often send-out tests with lengthy turnaround times, limiting their clinical utility. There is thus a critical need to explore diagnostic options that balance sensitivity, specificity, scalability, and breadth of coverage.

Previously, we described a novel rRNA-based strategy for rapid, sensitive bacterial identification directly from primary clinical samples or cultured blood samples (18) utilizing a commercially available assay platform for multiplexed hybridization-based RNA quantification (NanoString) (19). This assay, which we named phylogeny-informed rRNA-based strain identification (Phirst-ID), uses 192 probes targeting regions in the 16S and 23S rRNA from 98 medically relevant bacterial species (18). Each target region was selected for specificity at the level of a pathogenic species, or higher taxonomic levels. The fluorescent read-out from these probes results in differential hybridization that creates a unique “fingerprint” pattern of reactivity that accurately identifies each species. We later extended this approach to yeast, designing a Candida Phirst-ID probeset that discretely identified 11 medically relevant *Candida* species with 100% accuracy by targeting the 18 and 28S rRNA subunits (20). By recognizing the highly abundant rRNA itself, rather than the DNA that encodes it, this approach can detect and identify <30 *Candida* cells without amplification, as determined by serial dilutions (20), and the lack of enzymology in this hybridization-based assay enables it to work directly on crude lysates without nucleic acid purification (18, 20).

Here we further expanded this approach by creating a panel of 91 probes targeting 86 medically relevant fungi, targeting regions of the 18S and 28S rRNA subunits with differential conservation across taxonomic groupings, ranging from class to species (**Fig. 1**). The resulting “Pan-Fungal Phirst-ID” assay was first tested on a curated Training Set of 93 clinical isolates across 32 species of fungal pathogens, then verified on a Validation Set of 54 independent isolates. Using Pearson correlations, we achieved a positive species match rate of 84%. For some isolates, higher order taxonomic probes dominated the signature, compromising accuracy by overpowering more specific signals from genus and species level probes. To address this, we developed an optimized classifier, increasing species-level accuracy to 89%. To assess potential utility on primary specimens, we piloted the assay on archived clinical formalin-fixed paraffin-embedded (FFPE) tissue samples.

**Figure 1.**
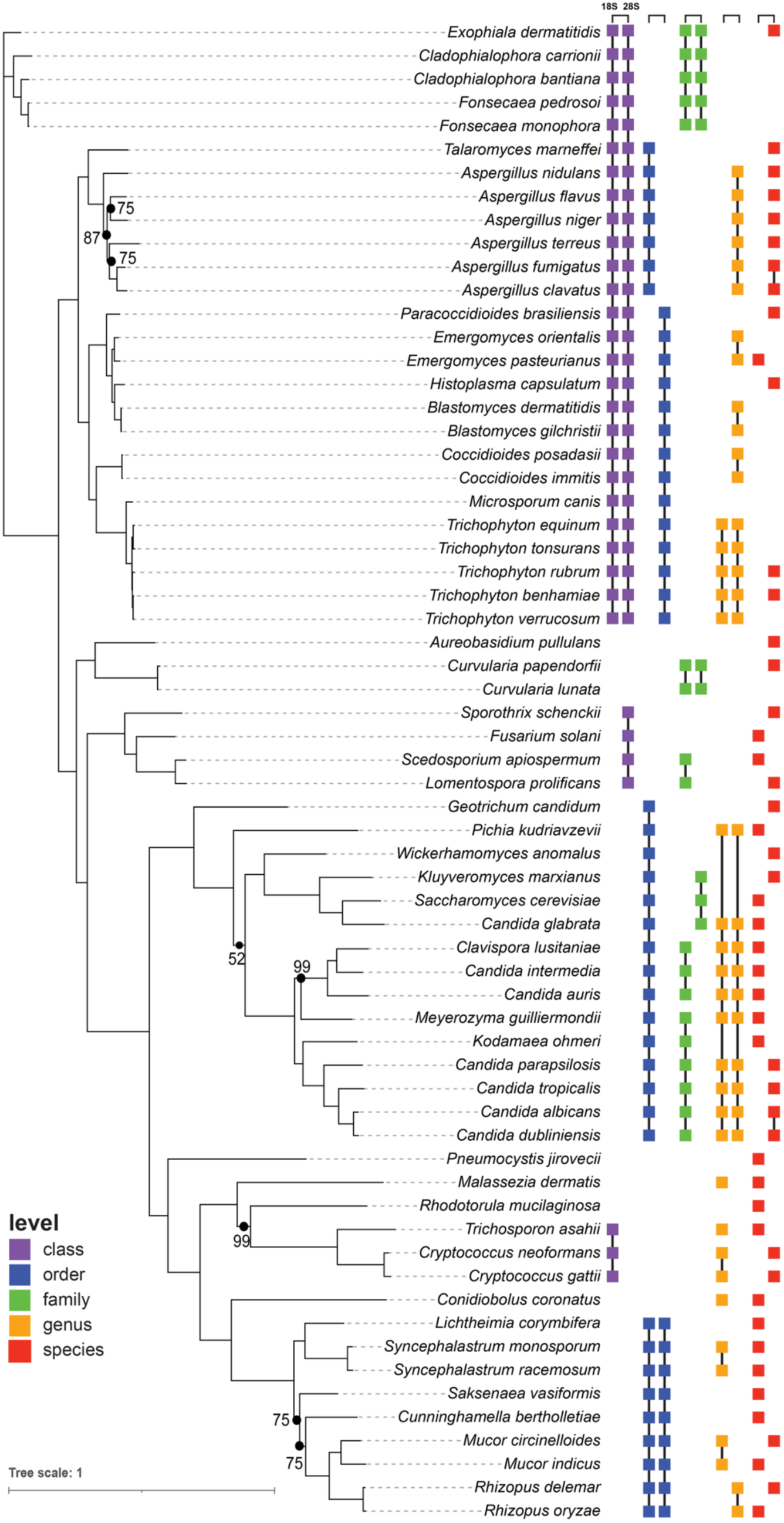
Phylogenetic coverage of Pan-Fungal Phirst-ID probeset. Fungal species included in probeset design, arranged by maximum-likelihood phylogeny inferred using IQ-TREE. To the right of each species, each solo square or set of connected squares denotes a unique probe; black lines connecting squares indicate a single probe designed to target multiple species within a taxonomic group. Colors indicate taxonomic level of the probes (see key at bottom left). For each level, squares at left indicate an 18S rRNA target and at right a 28S rRNA target. Tree branch lengths are in expected substitutions per site.

## Results

### Pathogen selection and Pan-Fungal Phirst-ID probeset design

We selected 86 medically relevant fungi using the WHO Fungal Priority Pathogen List (21) as key species to target with our panel, and an outgroup of 56 environmental and non-medically relevant fungi (**Table S1**). Targeting species-specific variable regions of the 18 and 28S rRNA subunits, we designed 60 species-specific probes (see Methods). For the remaining targeted species, we could not design a species-specific probe within the constraints of the NanoString assay due to phylogenetic constraints.

As previously described (18), we also designed higher-order taxonomic probes to improve accuracy in identifying target species through multiple recognition events, creating a unique probeset reactivity profile (PSRP) that serves as a “fingerprint” for each isolate tested. These higher taxonomic order probes can also enable partial characterization of pathogens not included in probeset design (18). To do so, we analyzed the 18S and 28S sequences for variable regions shared across species of the same genus, family, order, or class conserved enough to recognize all members within a given taxonomic classification, while excluding outgroup species (18, 20). This resulted in 14 genus probes, 8 family probes, 5 order probes and 4 class probes, culminating in a Pan-Fungal Phirst-ID probeset of 91 probes (**Fig. 1**; **Fig. S1, Table S2**).

### Collation of Training and Validation Sets

To rigorously test our panel, we collected isolates from 18 of 19 members of the WHO Fungal Priority Pathogen List, as well as 35 other clinically important species available from the MGH clinical microbiology laboratory. Isolates were split into Training and Validation Sets based on availability, with three isolates per species used for Training when possible and all remaining isolates used for Validation; when only three total isolates were available, two were used for Training and one for Validation (**Table S3a, b**; see Methods). Our Training Set contains 93 isolates spanning 18 unique genera and 26 defined species, and our Validation Set contains 54 isolates with the same genus and species numbers. For 7 genera, we were unable to get species characterization from the MGH clinical lab, due to standard clinical identification methodology. In addition, we tested 24 clinical isolates spanning another 14 genera and 21 species that were too rare (n = 1 or 2 in our collection) to include in our Training and Validation rubric, in an Underrepresented Set (**Fig. S2**). In all, we tested 171 isolates spanning 53 species across 32 genera, of which we can report accuracy statistics on 18 genera encompassing at least 32 unique species.

### Unique reactivity profiles accurately distinguish species

Pan-Fungal Phirst ID generated a unique PSRP for each species in the Training Set (**Fig. 2**) and the Underrepresented Set (**Fig. S2**), with reactivity primarily from the probes expected based on phylogeny of the tested isolate (purple boxes). To quantify differences in PSRPs between species, we first calculated a pairwise Pearson correlation coefficient for each sample against every other sample in the Training Set (**Fig. S3**). This simple analytical approach incorporates data from all probes tested in the probeset across the entire reactivity profile. Because of sequence conservation patterns in the targeted regions of rRNA genes, PSRPs should reflect species identity. As expected (18, 20), we observed the strongest correlations within isolates of the same species, whereas non-species matched pairs had lower correlation coefficients (**Fig. S3**). In a “leave-one-out” analysis for all isolates in the Training Set, 83% matched best to members of the same species, with most mismatches occurring to closely related species (**Fig. 3**). For 15 samples not identified to the species level, the best match in all 15 was to the same genus, the closest available taxonomic match in the Training Set. In all, 93% of isolates matched at the genus level, and match rates steadily increased for broader taxa, reaching 100% at the class level (**Fig. 3a**).

**Figure 2.**
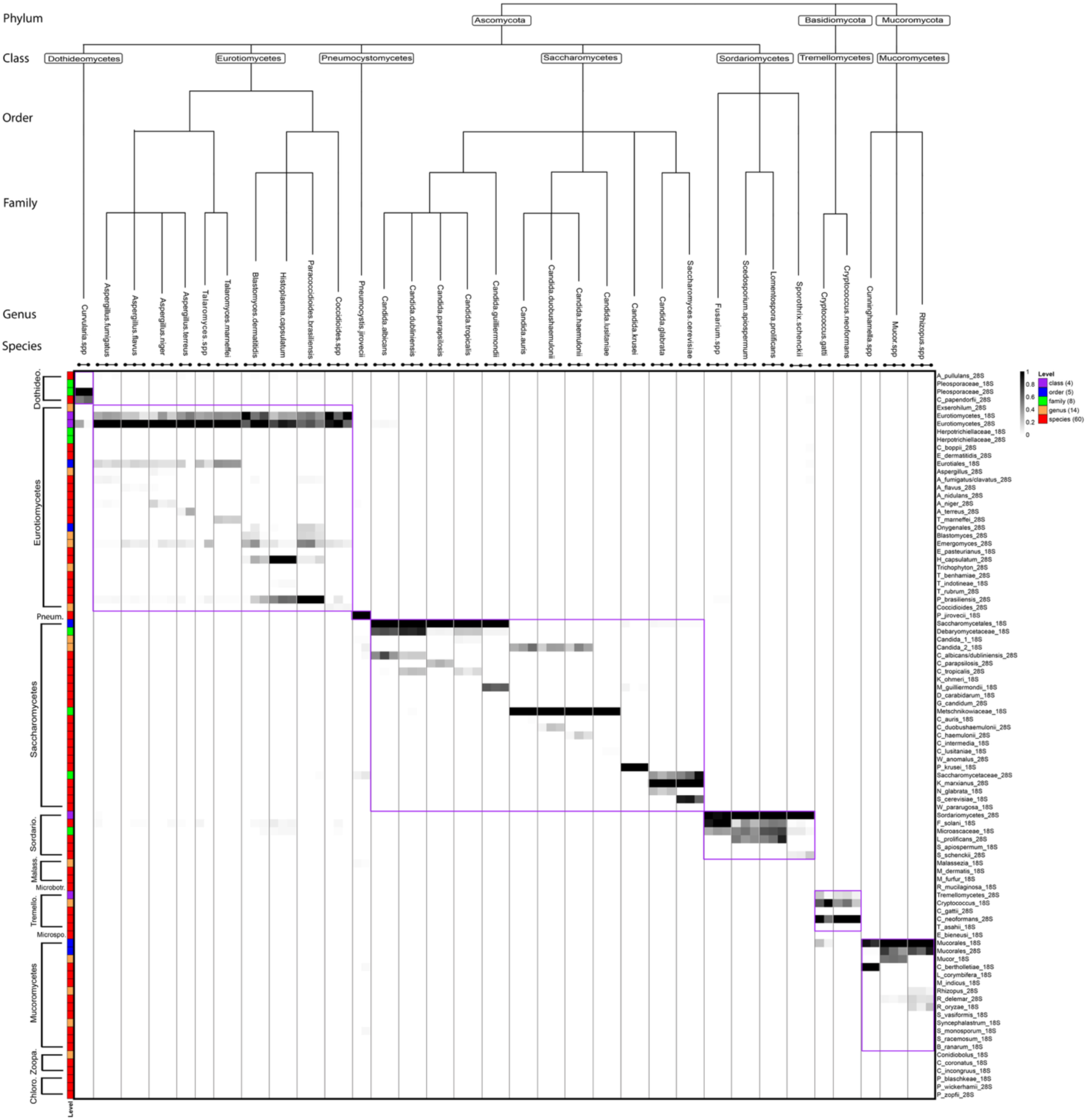
Pan-Fungal Phirst-ID panel generates unique PSRPs for 33 clinically relevant species. Heatmap shows scaled binding intensities of 91 probes (vertical axis) tested against the Training Set of 94 samples from 33 species (horizontal axis). Taxonomic level of each probe is indicated by colored box at left (numbers in parentheses = total probes designed at each level); probe name is listed at right. Each axis is ordered by taxonomic hierarchy, with species displayed in a cladogram. Horizontal black bars with dots indicate unique isolates of each species tested; vertical lines indicate transitions between species. Purple boxes indicate regions where the class of samples and intended probe targets match.

**Figure 3.**
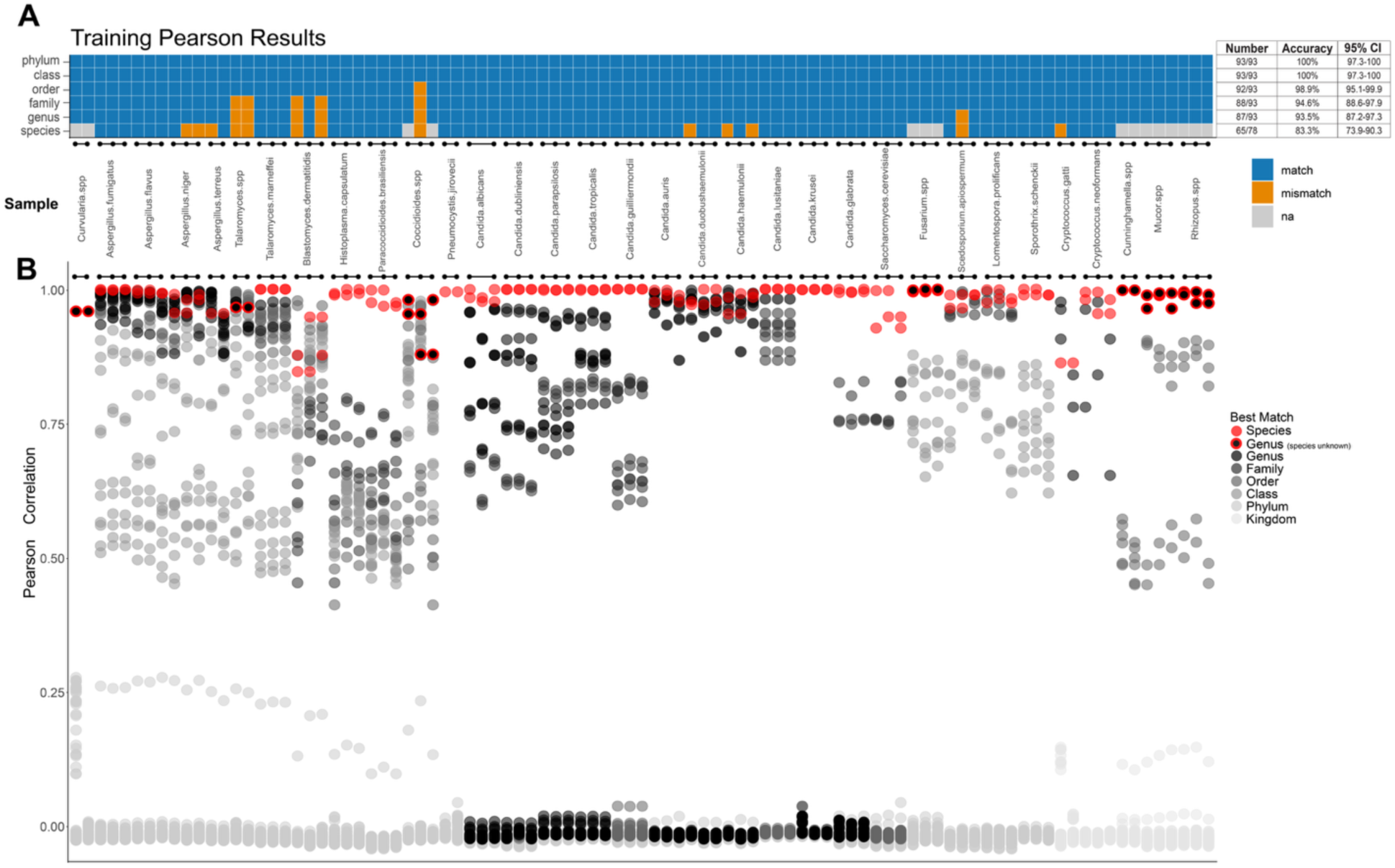
Pan-Fungal Phirst-ID PSRPs identify most species. Leave-one-out analysis of Training Set data showing closest taxonomic matches based on highest Pearson correlation. **(A)** Tile plot indicating accuracy of best non-self matches within the Training Set for identifying each individual isolate at each taxonomic level. **(B)** Dot plot of Pearson correlations of each indicated sample against all other samples in the Training Set, colored by closest-matched taxonomic level of the comparator. When the species of a test strain was unknown, the best possible match is to the genus level, indicated by a gray box at the species level in (A), and by a black circle with red border in (B).

### Leveraging taxonomy level information to improve classification accuracy

For many isolates in our training set, the higher taxonomic order probes resulted in greater signal than the species-specific probes (**Fig. 4a**). One notable example is the *Aspergillus* genus, where class probes dominated the signal readout compared with species probes (**Fig. 4b**). This systematic difference in probe binding by taxonomic level of the probe target, despite perfect complementarity between probe and target, was not observed in bacteria (18). This posed a unique challenge to accurate classification by Pearson correlation, which intrinsically weights higher signals and thus undervalues species-level probes with lower maximal reactivities, reducing species-level accuracy.

**Figure 4.**
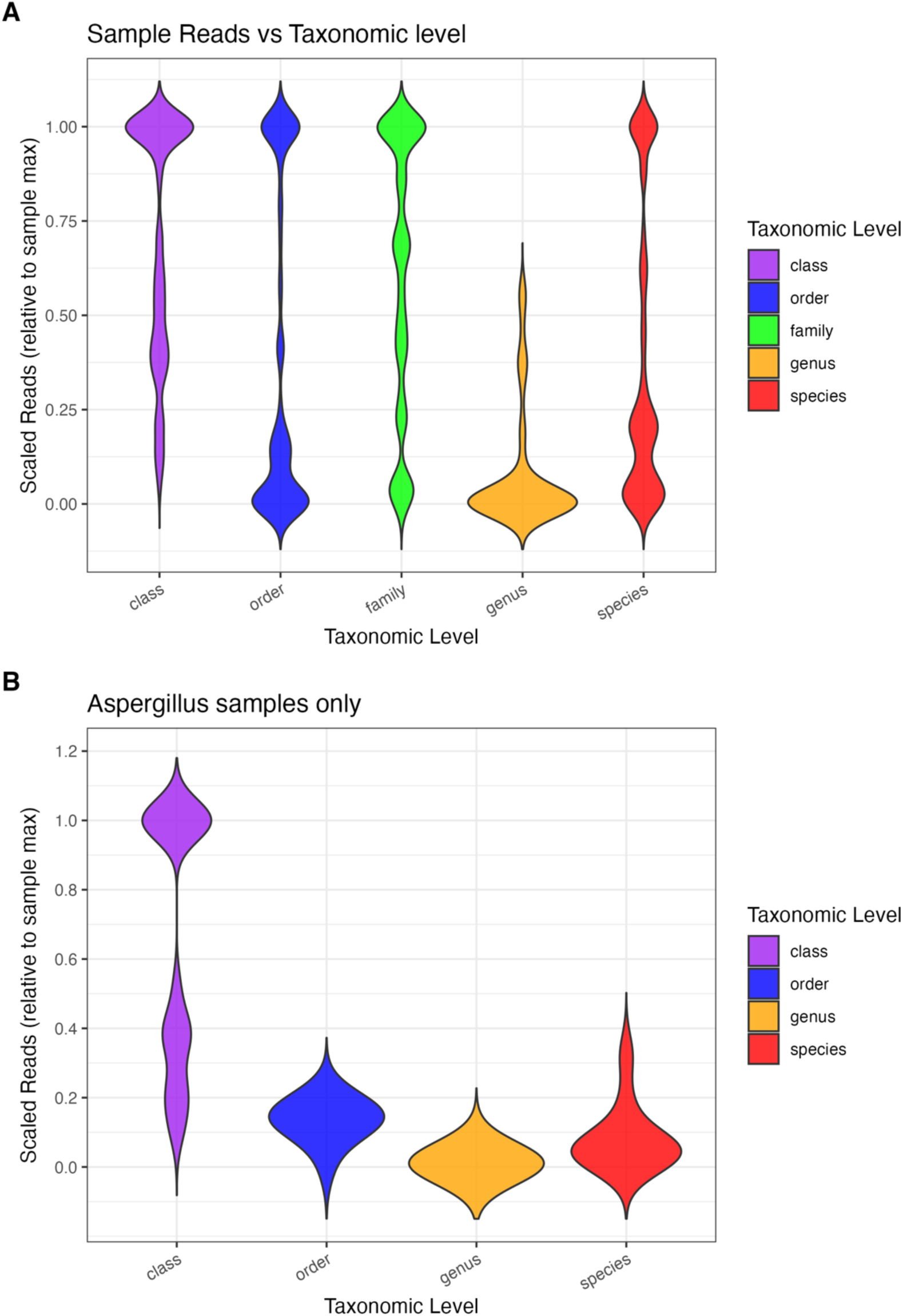
Probes designed for regions conserved at higher taxonomic levels exhibit higher binding signal. Violin plots of normalized signal intensity of probes targeting each taxonomic level of **(A)** all samples in Training Set and **(B)** only *Aspergillus* samples in Training Set. Y-axis for both graphs indicates non-zero probe reactivities normalized to the maximally reactive probe for each sample.

To address this uneven signal distribution across probes targeting different taxonomic levels, we developed a hierarchical classifier, still based on Pearson correlations but progressing stepwise through taxonomic classifications. We called this classifier **Co**mplementarity-based **P**hylogeny-**I**nformed **L**evel-**O**riented **T**raversal (Co-PILOT). This approach begins with the full PSRP, but traverses down the taxonomic classification, at each step eliminating any species and probes not included in the best-matched taxon. **Fig S4** illustrates a specific example of how Co-PILOT analyzes an *Aspergillus* sample, narrowing from all probes and samples (**Fig. S4a**), through those specific to the best-matched class (*Eurotiomycetes*), order (*Eurotiales*), family (*Aspergillaceae*), and genus (*Aspergillus*) (**Fig. S4b-e**). The best-matched sample from the Training Set at the final step, usually at the species level, is taken as Co-PILOT’s final identification. If the best match from the Training Set is a clinical sample only identified to the genus level (e.g., *Mucor* spp.), then Co-PILOT’s final step would be at the genus level. In this manner, the entire probeset is still used to classify each sample, but at each taxonomic level, only the probes designed to be informative in distinguishing among members of that taxon are considered. This method allows these more precise reactivity patterns to inform identity, rather than idiosyncratic differences between higher-order probes that might otherwise dominate a Pearson correlation. Critically, this novel classifier was devised and iteratively refined based only on the Training Set, before data from the Validation Set was examined. Thus, although Co-PILOT improved classification accuracy for the Training Set (**Fig. S5**), this should be viewed as “overtrained”. However, its performance on the Validation Set represents an independent assessment of classification accuracy.

### The Pan-Fungal Phirst-ID assay accurately classifies an independent Validation Set of diverse fungal pathogens

54 independent clinical isolates, representing the same genera and species as the Training Set, were tested as a Validation Set for the Pan-Fungal Phirst-ID assay (**Fig. S6**). The resulting PSRPs for these isolates were compiled into a separate dataset that was only released for analysis after the entire Training Set had been analyzed and the Co-PILOT algorithm had been finalized. Pearson correlations against each member of the Training Set identified a similar fraction of species (85%) and higher-order taxa in the Validation Set as in leave-one-out analysis within the Training Set (**Fig. 5a, b**). Co-PILOT performed better, identifying 89.3% of species in this independent Validation Set, with 96.2% concordance at the family level and >98% for all higher-order taxa (**Fig. 5c**).

**Figure 5:**
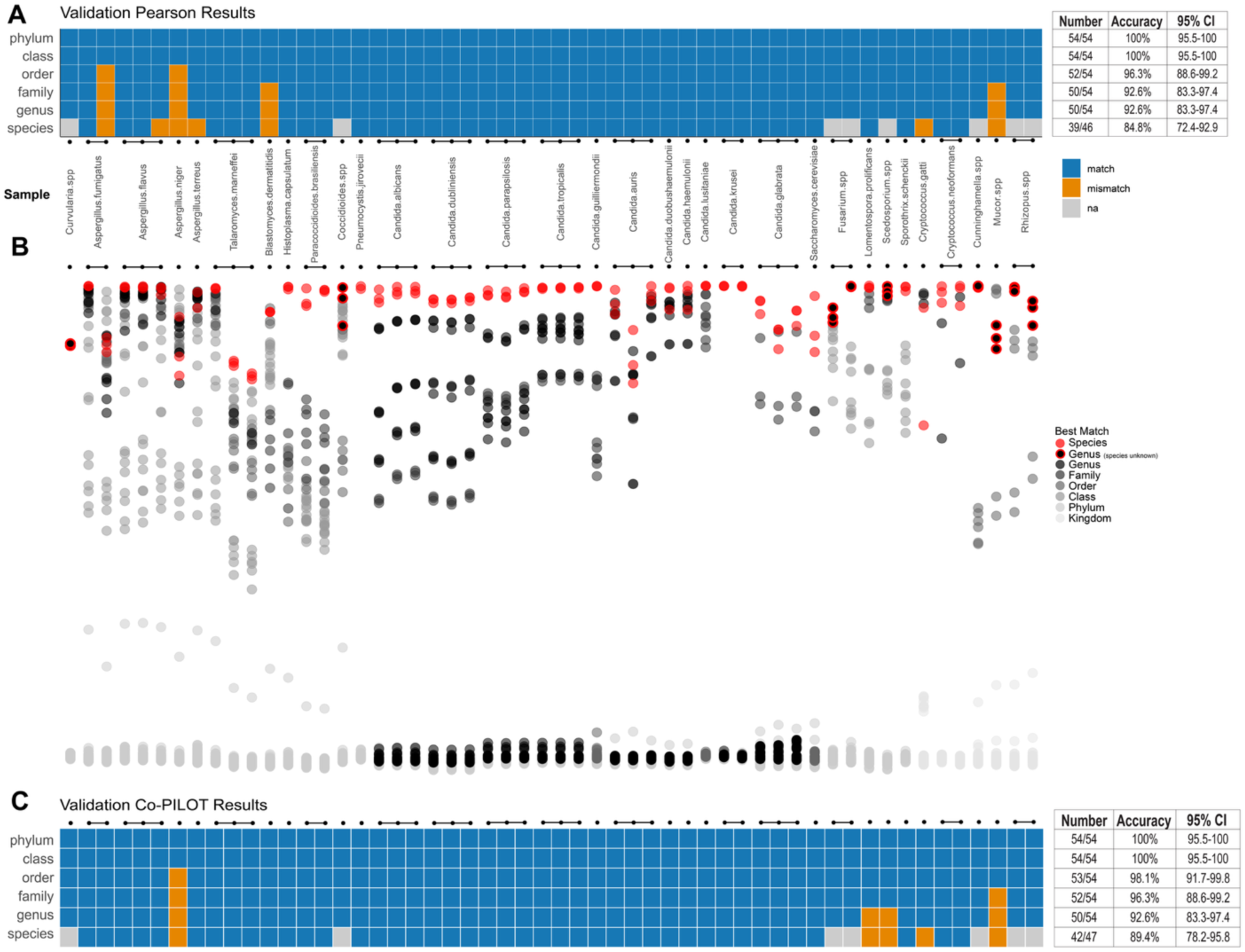
Pan-Fungal Phirst-ID PSRPs identify most species in independent Validation Set. Each sample in the Validation Set was compared by Pearson correlation with that of the entire Training Set. **(A)** Tile plot indicating accuracy of best matches at each taxonomic level based on highest Pearson correlation. **(B)** Dot plot of Pearson correlations of each Validation sample against all samples in the Training Set, colored by closest taxonomic level of the comparator. **(C)** Tile plot shows that Co-PILOT improves accuracy relative to simple Pearson correlations (A) for identifying the best match. When the species of a test strain was unknown, the best possible match is to the genus level, indicated by a gray tile at the species level in (A and C), and by a black circle with red border in (B).

To assess how the assay would perform when encountering an unknown sample outside the Training Set, we assessed Pearson correlations between the Training Set and the 24 samples from 21 species in the Underrepresented Set (**Fig. S7**). Whereas each tested sample from the Validation Set exhibited high Pearson correlations with its matched species in the Training Set (**Fig. 5b**), the highest Pearson correlation for each species in the Underrepresented Set varied widely (**Fig. S7**). Query samples with close phylogenetic matches in the Training Set exhibited high correlations, exemplified by *Candida* and *Aspergillus* species, but more distantly related samples such as *Prototheca* had highest correlations near 0.

### Investigation of discrepant classifications in the Validation Set

In the Validation Set, Co-PILOT misidentified 4 samples out of 54, all among species also misidentified in leave-one-out analysis in the Training Set. Incorrect ID matches were correctly binned at high order taxonomic levels but mismatched to different but related genera or species. Specifically, misclassifications were made between *Cryptococcus neoformans* and *Cryptococcus gattii*, and between *Scedosporium apiospermum* and *Lomentospora prolificans*. Both pairs are closely related phylogenetically; *C. gattii* was formerly referred to as a serovar of *C. neoformans* (22, 23), and *L. prolificans* was formerly classified as a *Scedosporium* (24). To assess the role of intrinsic assay variability in misclassifications, we examined correlations between assay replicates from the same isolate. Of 113 pairwise Pearson correlations across 57 isolates tested in replicate, 112 exhibited correlations > 0.94, with 80% > 0.99, suggesting that technical variability was unlikely to explain these misclassifications.

In examining reactivity profiles (**Fig. 2, Fig. S6**), despite probes designed to distinguish between each species pair, no real distinction exists between their PSRPs. For the pair of *Cryptococcus* species, a probe designed to selectively recognize the *C. gattii* 28S rRNA subunit failed to generate a signal for either species. This *C. gattii* species-specific probe matches a *C. deuterogattii* (VGII) sequence. Our training set included one *C. deuterogattii* sample, but this did not react with the species-specific probe, suggesting this probe has a general binding issue and should be redesigned and confirmed for alignment and potential reactivity to the other *C. gattii* species complex members. For *S. apiospermum* and *L. prolificans*, two problems arose in these closely related species: a probe designed to be specific for the *L. prolificans* 28S subunit cross-reacted with *S. apiospermum* as strongly as with *L. prolificans*; whereas a probe designed to selectively recognize the *S. apiospermum* 18S subunit generated no signal for either species. Relevant family and class probes recognized both species comparably, as expected.

We investigated one additional discrepant classification in the Validation Set, a sample clinically identified as *Mucor* that both Pearson correlations and Co-PILOT called a *Rhizopus*. On visual inspection, its reactivity pattern clearly resembled the *Rhizopus* from our Training Set more than the *Mucor* (**Fig. S8**). Given the difficulty in clinically distinguishing among the *Mucorales* by morphology (25, 26), we pursued ITS sequencing to clarify, which showed 100% match to *Rhizopus microsporus* (**Table S3b**), not a *Mucor* species, consistent with our assay’s call. By contrast, ITS sequencing confirmed the clinical identification of 2 other *Mucor* and 3 *Rhizopus* with concordant results (**Table S3a-b**). Reclassifying the discrepant isolate as a *Rhizopus* instead of a *Mucor* improves the accuracy of our Co-PILOT classifications on the Validation Set (to 91%, 94%, and 98% at the species, genus, and family levels respectively).

### Pilot use of Pan-Fungal Phirst-ID on clinical Formalin-Fixed Paraffin-Embedded (FFPE) tissue samples

Fixation of tissues using formalin and embedding in paraffin blocks is the gold standard method for preparing human tissues for histopathologic diagnosis, including microorganism identification (27–29). However, this process prevents subsequent growth of microbes; thus, if infection is only suspected after tissue fixation (e.g., in a lung nodule removed for suspicion of neoplasia), culture-based pathogen identification is precluded. Often such samples are obtained via surgical procedures that are impractical or even dangerous to repeat, meaning pathogen identification must be done either imprecisely by morphology, or via nucleic acid extraction and PCR from the fixed tissue (30, 31). Since Pan-Fungal Phirst-ID requires only hybridization and no enzymology, we hypothesized that it might perform comparably to PCR on crude nucleic acid preparations from FFPE tissue. We thus investigated the performance of our Pan-Fungal Phirst-ID probeset on nucleic acids extracted from archived clinical FFPE samples on which fungal forms were observed via microscopic examination, plus two negative controls with no fungal forms (**Fig. S9**).

Pan-Fungal Phirst-ID correctly identified some FFPE samples, largely depending on the quality of sample and NanoString read counts we were able to extract from each sample. When plotting heatmaps of read counts, 55% (15/27) displayed visually poor probe binding behavior; thus, prior to running Co-PILOT, we opted to exclude these as non-informative and focus our analysis on samples with sufficient NanoString reads (**Fig. S10**). Of these 12 remaining samples, 8 were correctly identified, with 2 others notably close: an *A. fumigatus* was called *A. clavatus*; and a *Madurella tropicana*, not previously tested, matched with the closest possible phylogenetic match, an environmental *Chaetomium* isolate (**Fig. 6**). We also found a negative control with a large *Candida albicans* signature, a possible contaminant in sample collection or processing; the other negative control showed low signal as expected (**Fig. S10**).

**Figure 6.**
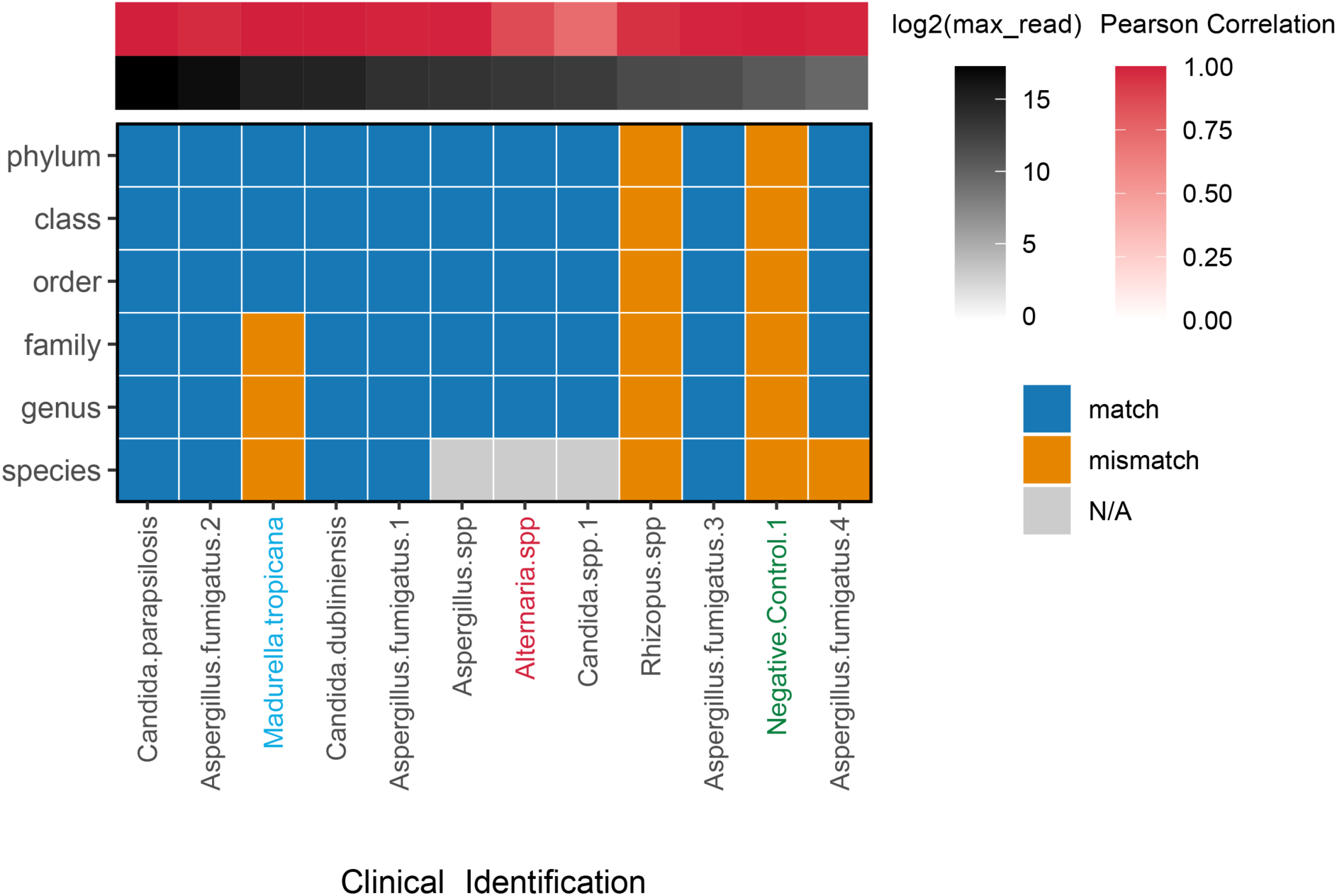
Pilot application of Pan-Fungal Phirst-ID to FFPE samples. Tile plot indicating prediction accuracy at each taxonomic level for the subset of FFPE samples with sufficient maximum probe reactivity to pass our threshold for Co-PILOT analysis. Sample order is based on the highest probe read count (black heatmap), which was used to set this threshold. Red heatmap shows Pearson correlation of each sample to its best match. Clinical laboratory identifications are listed below (see Table S3c); red text indicates an organism not included in probeset design, green text indicates a negative control FFPE sample (no fungal forms seen), blue text indicates an organism that was neither included in probeset design nor previously tested by Pan-Fungal Phirst-ID (and thus not available to the Co-PILOT algorithm for comparison).

## Discussion

Fungal infections are a growing global health problem, and current diagnostic methods are unable to meet the demands of healthcare systems. Currently available testing measures have significant limitations from financial burdens for instrument costs or expert personnel, frequent false positives for biomarkers, and limited organism range for which specific tests are available. We previously developed a molecular diagnostic approach based on rRNA hybridization, Phirst-ID, to rapidly and accurately identify bacterial pathogens (18) and *Candida* species (20) using a commercial detection system present in clinical pathology laboratories primarily for oncological use (32–34). Here, we extended this approach to target medically relevant fungi more broadly.

rRNA is an appealing target for hybridization-based identification due to its extreme conservation within species (35) and high abundance, allowing amplification-free detection. Experimental conditions for Phirst-ID only require base pairing, enabling detection from crude lysate with <30 minutes hands-on time and no nucleic acid extraction or purification, with a total assay time of <8 hours from a positive laboratory culture (18). By comparison, we previously reported a median of 28 hours from culture to identification for *Candida* bloodstream infections (20); other fungi take longer to identify. And whereas mass spectrometry-based methods must contend with surface proteomes that can vary across the fungal life cycle (5, 7, 9, 14), rRNA sequence is invariant.

Prior iterations of Phirst-ID identified organisms based on the closest Pearson correlation of PSRPs of a query organism to that of organisms in a reference panel (18, 20). By contrast, upon extending this approach to diverse fungal pathogens, we found that systematic variations in probe intensity across the Pan-Fungal Phirst-ID panel led to avoidable errors using this relatively simplistic classification scheme (**Fig. 4**). We therefore designed Co-PILOT, a classifier that integrates Pearson correlations of full and partial PSRPs with known taxonomic relationships between species to improve classification accuracy (**Fig. S4**). After iterative design on a Training Set, Co-PILOT improved identification on an independent Validation Set of 54 isolates compared with the simpler scheme.

Despite this, two pairwise misidentifications were made at the species level, each between close phylogenetic relatives: Co-PILOT mistook *C. neoformans* for *C. gattii* and vice-versa, and *S. apiospermum* for *L. prolificans* and vice-versa. Both errors appeared to be due to probes designed to distinguish each pair, not performing as expected. In the former case, the distinction between *C. neoformans* and *C. gattii* has minimal clinical consequences; though their geographic distribution is somewhat distinct, and their propensity to infect immunocompetent patients may somewhat differ, they both cause similar types of illness, with similar severity and management options (36). By contrast, the latter case is more clinically critical: though they cause a similar spectrum of illness in similarly immunocompromised patients, the intrinsic antifungal resistance profiles of *S. apiospermum* and *L. prolificans* differ meaningfully, leading to different empiric therapies (37, 38). To distinguish these, iterations on the probeset would be required, and if no rRNA-directed probe is able to make this clinically important distinction, mRNA targets may be needed, at a cost of assay sensitivity. Until these issues are fixed, this first iteration of the probeset requires that Phirst-ID reports of *Cryptococcus* should be considered to be either *C. neoformans* or *C. gattii*, and reports of *S. apiospermum* or *L. prolificans* should be considered to be either one, as the assay as currently configured is unable to distinguish these pairs.

In a pilot study to test additional clinical sample types on our Pan-Fungal Phirst-ID probeset, we tested archived FFPE tissue samples containing fungal forms. This sample type is critical for patient diagnosis when cultures are either not attempted or negative, or to identify species prior to culture, which for fungi can take days to weeks (1, 5, 6, 30, 31). FFPE tissue is optimized for morphological examination, but molecular methods are increasingly used to identify pathogens (31). Typical workflows involve DNA extraction followed by rDNA-directed PCR with amplicon sequencing, which generally has limited success, ranging from 13-34% in published series (39–43), typically worse for fungi (40). Of the 27 FFPE samples we tested in a pilot application, 12 passed our threshold of probe signal intensity (>1000 raw max probe counts); of these, Co-PILOT correctly identified 5 to the species level, 9 to the genus level, and one more to the closest available phylogenetic match (**Fig. 6**). For 14 samples positive for fungal forms by microscopy, we were unable to identify an organism due to lack of signal. Adding a bead beating step after deparaffinization helped increase signal for some samples, but more work is required to optimize RNA extraction for best results on this clinically important sample type, before testing head-to-head against the current standard of practice.

This study has several limitations. First, although Pan-Fungal Phirst-ID was designed using sequences from 86 fungal pathogens, we have thus far only been able to rigorously test and independently validate it against multiple isolates of 32 species. Its accuracy in identifying these 32 key species should be well predicted by its performance on our Validation Set and should improve as the Training Set gets larger with further use. However, its performance on species not included in this Training Set is uncertain. We did observe predictable recognition patterns on an additional 21 species (**Fig. S2**) from our Underrepresented Set. For these species not included in the Training Set, Pearson correlations to the Training Set were highest for those with a genus-level match (**Fig. S7**, see *Candida* and *Aspergillus spp.*). When trained on sufficient examples of new species, Pan-Fungal Phirst-ID should perform well, as long as the 91 probes provide a unique PSRP. However, recognizing when an off-panel species is encountered remains a key challenge, and care will be needed in results reporting, particularly for unknown samples that show low correlations to Training Set isolates. Second, although Pan-Fungal Phirst-ID with Co-PILOT accurately identified >90% of clinical isolates samples at the species level and 98% at the family level in an independent Validation Set, it clearly struggled with some clinically relevant distinctions discussed above, most notably between *S. apiospermum* and *L. prolificans*. Third, with a maximum of 3 samples each in the Training and Validation Sets, assay performance across phylogeny remains unexplored, though the extreme conservation of the rRNA operon suggests less impact of diversity on this assay than other targeted assays. Additionally, this high conservation also comes with costs, including difficulty distinguishing some closely-related species. Fourth, several isolates in our Training and Validation Set were only identified to the genus level by the clinical microbiology laboratory, and therefore Phirst-ID accuracy could only be measured to the genus level. With further training, it is possible that species-level distinctions could be made, but genus-level identification for these isolates meets or even exceeds current clinical laboratory reporting standards for these genera (e.g., *Mucor*, *Rhizopus*, and *Cunninghamella* are often reported as order *Mucorales* from clinical laboratories, whereas Pan-Fungal Phirst-ID accurately identified them to the genus level - better than clinical morphology-based assessments in at least one case; **Fig. S8**). Fifth, aside from the pilot on FFPE tissue, we thus far only trained and tested this assay on monocultures. Although *Candida* Phirst-ID was able to recognize mixed cultures as linear combinations of PSRPs from each species (20), the increased complexity of PSRPs in the Pan-Fungal probeset will likely make this more challenging. Sixth, in this initial work, we rigorously tested this assay primarily on cultured isolates, although the hybridization-based assay in other contexts has worked well despite an excess of off-target nucleic acids (such as human tissue). Still, despite some promising results on a small number of pilot samples, further optimization is needed for deployment on primary clinical samples, especially the high-value clinical use case of FFPE tissue, to optimize sensitivity and diagnostic yield. Finally, this assay is currently research-use-only, like its bacterial and *Candida* counterparts.

Though it was designed with clinical use in mind, this broad-range, sensitive assay could be readily developed for other diagnostic settings, including environmental, agricultural, or biodefense monitoring. We demonstrate the potential for design across a wide range of species, and it would be feasible to combine our bacterial probeset (18) with this Pan-Fungal probeset to characterize complex samples or polymicrobial infection (20). Additional work beyond the scope of our academic laboratory would be required to optimize process and reagents, and ideally automate portions of the workflow, prior to implementation and clinical validation. Once optimized and scaled, Pan-Fungal Phirst-ID offers a novel approach to fungal identification that combines features of the non-enzymatic workflow of mass spectrometry with the sequence conservation and high-abundance targets of rRNA-based methods, implementable on either cultured isolates or primary specimens.

## Materials and Methods

### Probeset Design

Guided by the WHO Fungal Priority Pathogen list, we compiled 18S and partial 28S sequences for 86 medically relevant fungal species from Silva (44) or NCBI databases to train and validate our probeset. Additional 18S sequences were curated for 56 environmental fungal species to serve as an outgroup during probe design to minimize unintentional cross reactivity (**Fig. S1, Table S1**; See Supplemental Methods for sequence sources and rationale for taxon-level probe design). Employing a similar strategy used previously (18), we profiled each desired target sequence using NanoString’s proprietary probe design algorithm to identify all putative pairs of consecutive 50mer probe-binding regions that meet parameter specifications. Specifically, probe sequences are assessed for predicted binding kinetics, thermodynamics, secondary structure, and sequence composition for the desired targets while minimizing predicted cross-reactivity against non-targeted outgroup species, or the human genome. Probes were designed either to bind exclusively to a single species (species-specific probes) or to bind all members of a higher taxon while excluding non-members (genus, family, order, and class level probes). For *Candida* species, which have undergone considerable phylogenetic reclassification of late, we kept the clinically familiar genus classification *Candida* for all (*Candida, Candidozyma, Pichia, Nakeseomyces, Meyerozyma*, etc.), which likely changed design constraints on the genus-level probe. This decision could be revisited in future iterations of the probeset.

The final probeset includes 60 probes designed to be species-specific, as well as 31 probes designed to recognize rRNA regions conserved across higher taxonomic groupings from genus to class (**Fig. S1**). Distinguishing the closely related *Candida albicans* and *Candida dubliniensis,* and *Aspergillus fumigatus* and *Aspergillus clavatus*, was not possible at the species level, so we designed single probes predicted to selectively recognize each pair of species. During testing, the *Mucor circinelloides* 28S probe yielded substantial cross-reactivity against nearly all tested species and was thus removed from the final probeset (**Table S1**). Also included in the probeset are 12 control probe pairs (6 negative, 6 positive) directed at External RNA Controls Consortium (ERCC) targets as previously described (18).

### Sample acquisition

110 clinical isolates across 47 species were collected from the Massachusetts General Hospital (MGH) Clinical Microbiology Laboratory. Isolates were identified through standard clinical microbiology workflows as part of routine patient care and shared with the investigators under Mass General Brigham Institutional Review Board protocol 2015P002215. In keeping with diagnostic limitations in these standard clinical workflows, N isolates from 7 genera were only identified to the genus level. Another 35 isolates from 11 species were sourced from strain repositories (the United States Centers for Disease Control’s Antimicrobial Resistant Isolate Bank, or the American Type Culture Collection). 26 isolates across 7 species were obtained from external collaborators to expand species coverage, particularly for organisms not routinely cultured in our laboratory. Per our biosafety regulations, all cultured isolates were received in Trizol. For 6 species for which we were unable to obtain a cultured isolate, we used either gDNA or RNA as available. Coded FFPE tissue samples (n = 27) spanning a range of fungal burdens were obtained from the Brigham & Women’s Hospital Pathology Department (2019–2025). A full list of samples, sources, and identification methods is provided in Table S3 and Supplemental Methods. All fungal isolates obtained were tested and included in this study.

### Training and Validation Set parameters

To test the Pan-Fungal Phirst-ID probeset, we acquired as many samples from our fungal pathogen list as possible. In all, we collected and tested 171 samples (isolates, gDNA, or RNA) from 53 species. To rigorously assess assay performance, we divided these samples into Training and Validation Sets based on the number of samples of each species we were able to collect (**Table S3a, b**). In the ideal case, 3 samples of the same genus and species were included in the Training Set. Any remaining samples (up to 3 more) were assigned to the Validation Set. If we were only able to obtain 3 samples total, we included 2 in the Training Set and assigned third to the Validation Set (**Table S3a, b**). These parameters resulted in a Training Set containing 93 samples across 32 species and a Validation Set containing 54 samples across the same 32 species. For 21 additional species, we were only able to obtain 1 (n = 18) or 2 (n = 3) samples. Since this was insufficient to assess classification accuracy, we report the results of this Underrepresented Set (**Table S3c**) separately. For assessment of classification performance, background subtraction and normalization was carried out exactly as previously described (18, 20), and classifiers were developed and iterated using only data from the Training Set, before being run on the Validation Set with no changes to any analytical processes. All Validation Set analyses were done with authors blinded to sample identification until after predictions were finalized. For FFPE samples, Pathology co-authors (MLD-M and IHS) selected samples, whereas the remaining co-authors (notably EAY, BJB, CAC, RPB) stayed blinded to the identities of all species until all thresholding decisions, classification algorithms, and identity predictions were finalized. In all cases, accuracy is reported based on concordance with standard identification methods, either from standard clinical microbiology laboratory workflows for the 86 clinical isolates, or by report of the providing laboratory (typically from whole genome sequencing, or from original clinical identification) for the 61 isolates obtained from research laboratories or strain repositories. Confidence intervals for accuracy statistics are computed via Jeffreys method (45).

### Sample processing

#### Cultured isolates collected from MGH

125μL of each sample in Trizol was added to a 2ml screw cap tube (MP Biomedicals, CAT# 76044-692) with an additional 125μL of Trizol reagent (ThermoFisher CAT#15596026) and 100μl of 0.1μm zirconia/silica beads (Biospec, CAT# 11079101z), then bead-beaten for 2 rounds on the FastPrep 5G (MP Biosciences) for 90s, at 10 m/s. 50μL of chloroform was added to each sample, inverted, and the mixture was spun down at 12,000 x g for 15 minutes at 4C. 50μL of the aqueous phase was transferred to a fresh 1.5 ml Eppendorf tube. To prevent degradation of the RNA in this crude lysate, 50μL RLT buffer (Qiagen CAT# 79216) with 1% β-mercaptoethanol (β-ME), an irreversible chemical RNase inhibitor, was added to each lysate prior to storage at -80°C.

#### gDNA samples of selected isolates

For gDNA samples obtained from the Fungal Genomics group at the Broad Institute, approximate concentrations were measured using the Nanodrop One system (ThermoFisher CAT#ND-ONE-W). Dilutions of some gDNA samples were made in 1x Phosphate Buffered Saline (PBS) to ensure probe reactivity patterns were within control cutoffs and max probe limits as described below.

#### Cultured isolates prepared from ARBank isolates

The ARBank *C. auris* (CAU) and drug-resistant *Candida* (CAN) collections were used to supplement *Candida* species for Training and Validation sets (**Table S3a, b**). Cultured isolates were prepared in house and processed as previously described (20).

### NanoString Data generation

Our custom Pan-Fungal Phirst-ID probeset was run using the Elements assay variation on the standard NanoString assay for multiplexed RNA detection. Briefly, lysates from cultured organisms were diluted 1:10 in PBS to avoid assay saturation. 1.5μL of lysate or nucleic acid was incubated with unlabeled probe pairs for each target (IDT) and Elements TagSet-96 reagents (NanoString CAT#121000608). Hybridization conditions were standard, except for 2 modifications: lysates were incubated at 95 °C for 2 minutes immediately prior to hybridization to denature secondary structural elements and disrupt protein binding in rRNA targets, and hybridizations were incubated for one hour instead of the recommended 16-24 hours, exactly as we have previously done (18, 20). Hybridized samples were then run on the NanoString nCounter platform and subsequently analyzed using the nCounter Digital Analyzer.

Cell densities from samples cultured by collaborators, and from paraffin scrolls, were unknown prior to running on the nCounter platform. We established three parameters to address sample overloading: (1) positive control signals out of the expected order given known spike-in concentrations, suggesting cross-reactivity; (2) any negative control value > 100 total read counts; or (3) maximum probe count >100,000, where saturation might affect relative ratios. If any of these 3 parameters were met, the affected sample was re-run with increased dilutions until the issue resolved. The maximum number of re-runs required for a given sample was up to three to meet our stated parameters. Across Training and Validation Sets, 62 lysates required at least one re-run, with four samples requiring additional runs. All such decisions were made solely based on these three criteria, prior to any assessment of prediction accuracy.

### Data processing and visualization

Raw binding data (counts per probe) were compiled using NanoString nSolver software (v3.0). Raw counts were normalized using positive and negative control spike-ins provided by NanoString using custom scripts in R, and background signal for each probe was subtracted based on the average of three blank lanes, exactly as previously described (18). Pearson correlations for the resulting normalized, background-subtracted probeset reactivity profiles were subsequently calculated across samples using the “cor” function in R (version 4.3.1). Heatmaps were generated by the Pheatmap package v1.0.12 in R. Probes and samples were arranged phylogenetically unless stated otherwise.

### Co-PILOT Classification

We developed a hierarchical classification model based on Pearson correlations (**Fig. S4**) termed **Co**mplementarity-based **P**hylogeny-**I**nformed **L**evel-**O**riented **T**raversal (Co-PILOT), coded entirely in R (https://github.com/broadinstitute/FungalDx_ID/). Initial Pearson correlations were calculated for each isolate to identify most similar samples based on probe-binding signatures. The taxonomic group (starting with class) was then found for the closest match. All other isolates not in this taxonomic group were then removed, as were probes not designed to recognize that group (**Fig. S4a-b**). This process was repeated until the species level, or if there were no more samples to compare to. The Co-PILOT algorithm was developed and iterated using only our Training Set, then deployed without modification on our Validation Set, considering each Validation sample one at a time and using matches to the Training Set as its basis for identification decisions. When the best match was to a sample from the MGH clinical laboratory for which identification was only determined to the genus level, Co-PILOT was unable to make a species prediction but could predict identity to the genus level.

### Pilot study of FFPE samples

FFPE scrolls (60μm per block in each tube) were obtained from the Brigham and Women’s Hospital Pathology Department. Samples FP001-FP012, FP016-019 were processed using the RNeasy FFPE kit (Qiagen CAT# 73504), per NanoString recommendations. Given inconsistent results at an interim assessment, we processed the remaining samples (FP020-030) using the Quick-DNA/RNA FFPE kit (Zymo CAT# R1009) (**Table S3c**). Samples were first deparaffinized using proprietary solutions provided by each kit, then heated to either 56°C followed by 80°C (Qiagen) or 54°C (Zymo), separating wax from tissue. Samples underwent two rounds of bead beating as described previously and spun down at 13,000xg for 1 min before loading onto column for final purification. All lysates were stored at -80°C after elution. FFPE samples were analyzed as a separate dataset relative to Training and Validation Sets. Compiled FFPE data was analyzed for species classification against a “superset” containing Training, Validation, and Underrepresented Sets, mirroring how we would expect to deploy this method on primary tissue samples going forward. After visual inspection of the resulting data for all samples, we noted that 14 samples with visible fungal forms had <1000 raw counts from any probe and failed to show distinctive reactivity patterns (see **Fig. S9**). Using this metric as a threshold for usable signal, we divided samples based on their max probe counts into groups that had sufficient (n = 12) or insufficient (n = 15, including one negative control) reads, though we report Co-PILOT’s best match for each sample in each group. As described above, if a species identification was not defined for a given sample, the final best match was made at the genus level.

### ITS sequencing workflow to confirm species ID

Clinical isolates were received in Trizol, and additional cultured material was unavailable. For those unable to be identified to the species level by standard workflows, we sought to obtain genomic DNA (gDNA) to enable ITS sequencing. As Trizol facilitates isolation of RNA rather than DNA, we omitted the chloroform extraction, which enhances partitioning of RNA from gDNA, to improve gDNA recovery for PCR. 100μL of sample was combined with an additional 300μL of Trizol, underwent 2 rounds of bead beating as previously described, then was processed using the Zymo DirectZol RNA kit (CAT#: R2050). ITS1 and ITS2 regions were amplified using the NEB Q5 hot start master mix (CAT#:M0491S) using primers and cycling conditions as previously described (20, 46). Samples with bands corresponding to either ITS1, ITS2 or both on gel electrophoresis were purified using the Zymo DNA Clean and Concentrate (CAT#: D4033) and sent to Plasmidsaurus for amplicon sequencing. Results were first analyzed in Geneious and trimmed for quality using standard parameters; trimmed sequences, then were analyzed via UNITE (47) or NCBI BLAST to identify the best-matched species.

## Data Availability

All data produced in the present study are available upon reasonable request to the authors

https://github.com/broadinstitute/FungalDx_ID

## Acknowledgments

We thank Dr. Sinem Beyhan and Jacob Durazo from the J. Craig Venter Institute, and Dr. Joseph Kovacs, Dr. Shelly Curran, and Dr. Liang Ma from the NIH, for generously sharing samples, and Andrew Grootsky, Aleksandr Kutchma, Richard Boykin (current or former employees of NanoString, now Bruker Spatial Biology) for their assistance with our non-standard probe design. This work was supported in part by the National Institutes of Health [grant numbers 1R01AI153405 to R.P.B. and C.A.C., and T32 AI007061 to E.A.Y]. The content is solely the responsibility of the authors and does not necessarily represent the official views of the National Institutes of Health. The funders had no role in study design, data collection, and interpretation, or decision to submit this work for publication. R.P.B, E.A.Y and M.L.D-M are co-inventors of subject matter disclosed and claimed in U.S. Patent No. 19/631,821, titled “Compositions and methods for rapid RNA-based detection of fungi”, co-owned by Massachusetts General Hospital and The Broad Institute, Inc., directed to methodology of identifying fungal pathogens using rRNA and disclosure of fungal probes, as described in this manuscript.

