## Supplementary Information (Methods and Figures S1-S10) for "A rRNA hybridization-based approach for rapid and accurate identification of diverse fungal pathogens"

[1]Infectious Disease and Microbiome Program, Broad Institute of MIT and Harvard, Cambridge MA 02142 USA
[2]Genomic Center for Infectious Disease, Broad Institute of MIT and Harvard, Cambridge MA 02142 USA
[3]Present address: Valo Health, 75 Hayden Avenue Suite 200, Lexington, MA 02421 USA
[4]Department of Pathology, Massachusetts General Brigham, Harvard Medical School, Boston, MA USA
[5]Department of Pathology, Brigham and Women's Hospital, Harvard Medical School, Boston, MA 02115 USA
[6]Department of Molecular Microbiology and Immunology, Brown University, Providence RI 02912 USA
[7]Microbiology Laboratory, Department of Pathology, Massachusetts General Hospital, Boston, MA 02114, USA
[8]Infectious Diseases Division, Department of Medicine, Massachusetts General Hospital, Boston MA 02114 USA
*Denotes Co-First authorship: EAY and BJB contributed equally.
[#]To whom correspondence should be addressed

### Supplementary Information: Contents

p. 1 Index
p. 2-3 Supplementary Methods
p. 4-14 Supplementary Figures S1-S10
p. 15-16 Supplementary References
(note: Supplementary Tables available as a separate file)

### Supplementary Methods

#### Sample Acquisition

Isolates only able to be identified to genus level in the clinical microbiology laboratory included all *Curvularia*, *Coccidioides*, *Fusarium*, *Cunninghamella*, *Mucor*, *Rhizopus* and *Talaromyces* isolates. Outside of samples acquired from our own laboratory and the ARBank, additional samples were generously shared with us from the following sources: *Histoplasma capsulatum* (n = 3) cell pellets suspended in Trizol from Dr. Sinem Beyhan (JCVI) (1); *Pneumocystis jirovecii* gDNA from human autopsy samples from Dr. Joseph Kovacs (NIH) (n = 3); and *Blastomyces dermatitidis* total RNA, as well as gDNA from *Cryptococcus gattii, Paracoccidioides brasiliensis, Sporothrix schenckii* and *Talaromyces marneffei* from the Fungal Genomics Group at the Broad Institute (n = 19) (1–5).

Coded/de-identified FFPE tissue samples (n = 27) were obtained from the Brigham & Women's Hospital (BWH) Pathology Department, after visual screening to identify archived clinical samples from 2019-2025 with varying levels of visible fungal forms (ranging from rare to confluent). Fungal identification of these autopsy and surgical pathology cases were based on integration of all available laboratory results from anatomic pathology histochemical stains, culture isolation from concurrently collected tissue samples, and molecular testing of FFPE tissue, frozen tissue, or culture isolates (6). Supplementary Table S3d contains additional information on each FFPE sample.

#### Probe Design

**Reference Sequences:** 18S and partial 28S sequences for the 86 ingroup species were primarily obtained from the Silva database (7). Where Silva sequences for target species were too short, or unavailable, regions were retrieved from NCBI assemblies for the following species: *Mucor circinelloides*, *Emergomyces orientalis*, *Epidermophyton floccosum*, *Coccidioides posadasii*, *Coccidioides immitis*, *Histoplasma capsulatum*, *Emergomyces pasteurianus*, *Exophiala dermatitidis*, *Trichophyton indotiniae*, *Kodamaea ohmeri*, *Wickerhamomyces anomalus*, *Kluyveromyces marxianus*, *Malassezia dermatis*, *Enterocytozoon bieneusi*. All retrieved sequences were validated using an alignment of each gene and manual inspection. While 18S sequences appeared largely complete, due to inconsistent annotation of 28S sequences, full-length 28S sequences could not be obtained for all species.

**Design rationale for species and taxon-level probes:** For species-specific probe design, an outgroup of all other pathogenic and environmental fungi was provided to optimize specificity (8). This approach aimed to design probes predicted to bind uniquely to the targeted species in a given rRNA region (Table S2). Probes were also designed to uniquely target higher-level taxa (genus, family, order, or class), including all members of that taxon but no non-members, across both pathogens and environmental fungi. The inclusion of non-pathogens provided necessary sequence diversity to identify taxonomic probes.

**Control probes:** The 6 positive control probes target External RNA Controls Consortium (ERCC) spike-ins included in every hybridization reaction at pre-specified concentrations; the 6 negative control probes target bioorthogonal sequences not present in the hybridization. All control probes are provided as part of standard NanoString hybridizations (9).

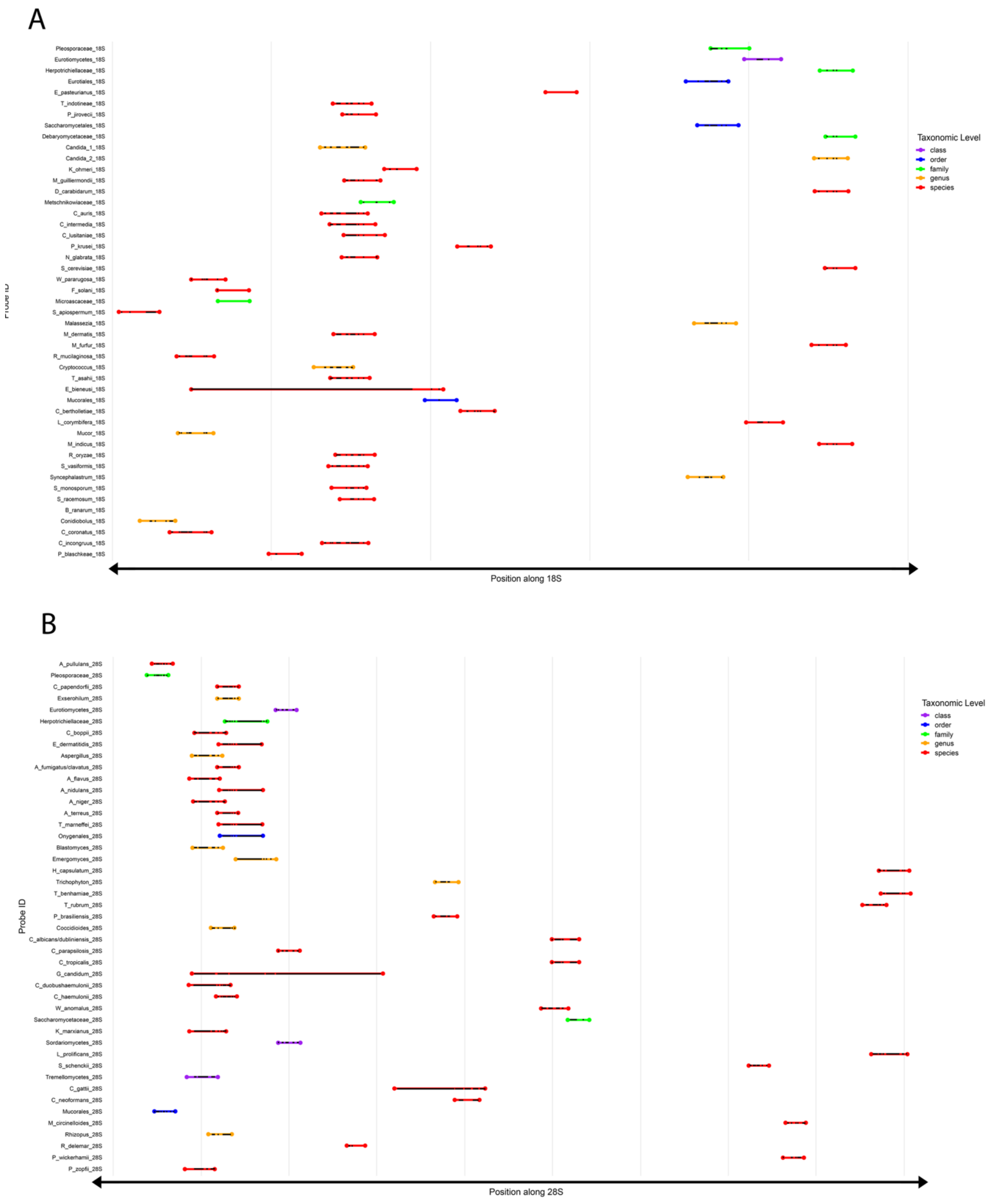


**Figure S1. Coverage maps of Pan-Fungal Phirst-ID probe sequences along 18S and 28S rRNA subunits.** Probe sequences mapped along target rRNA sequence of (A) 18S and (B) 28S subunits. Color of line indicates taxonomic level specificity of probe. Longer mappings indicate gaps in alignment due to genetic variation in the consensus sequence. Light gray vertical lines indicate 500 bp distance along the consensus 18/28S axes (including gaps). Probe sequences are available in Supplementary Table S2.

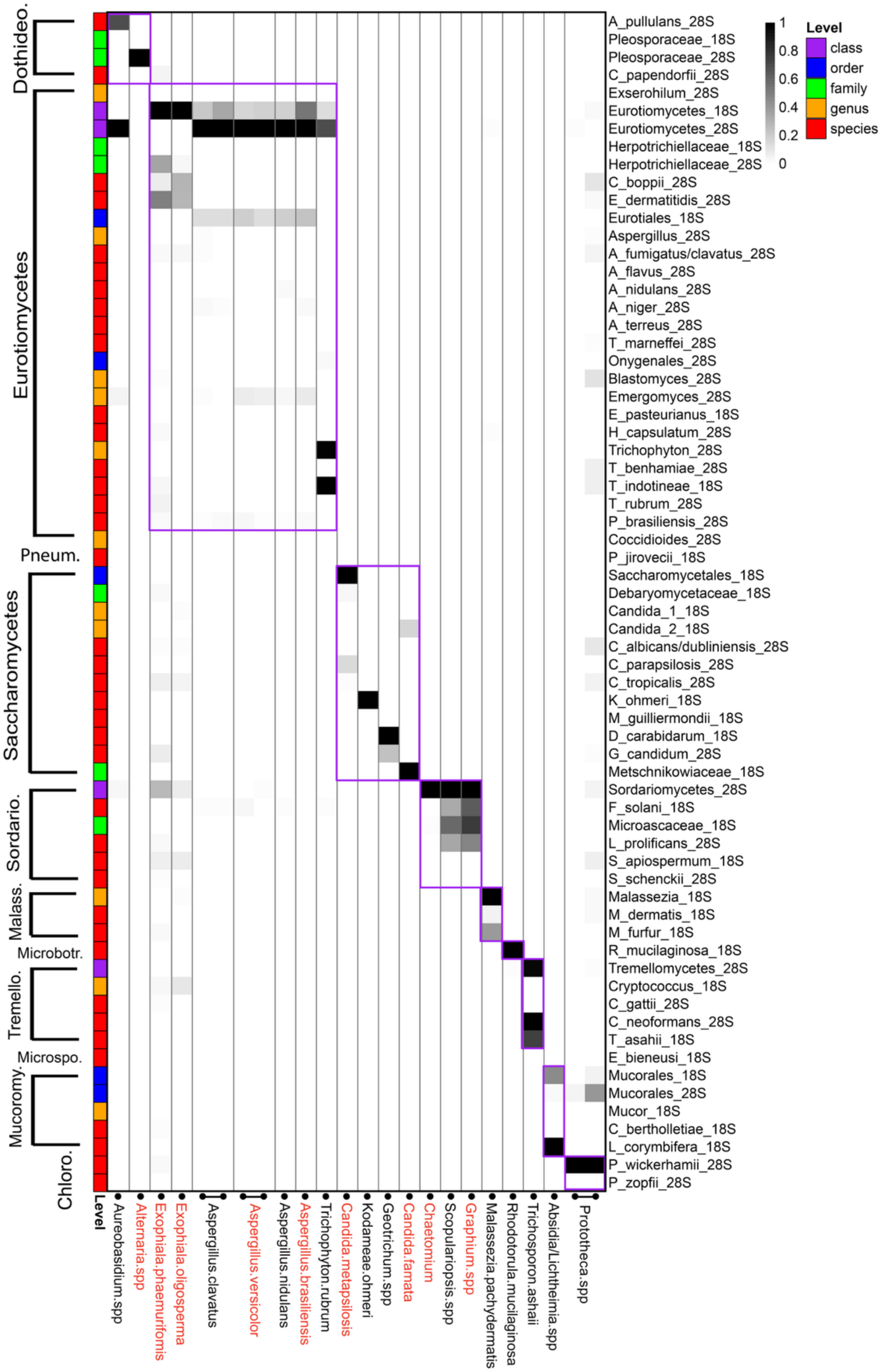


**Figure S2 Pan-Fungal Phirst-ID PSRPs from the Underrepresented Set.** Heatmap displaying normalized probe binding intensities of 24 additional samples, for which we were only able to obtain 1 or 2 examples and thus did not meet criteria for inclusion in the Training or Validation Sets (because we could not rigorously apply our sample identification algorithms). Purple boxes indicate regions where the class of samples and intended probe targets match. Red text indicates species not included in probeset design but obtained for testing.

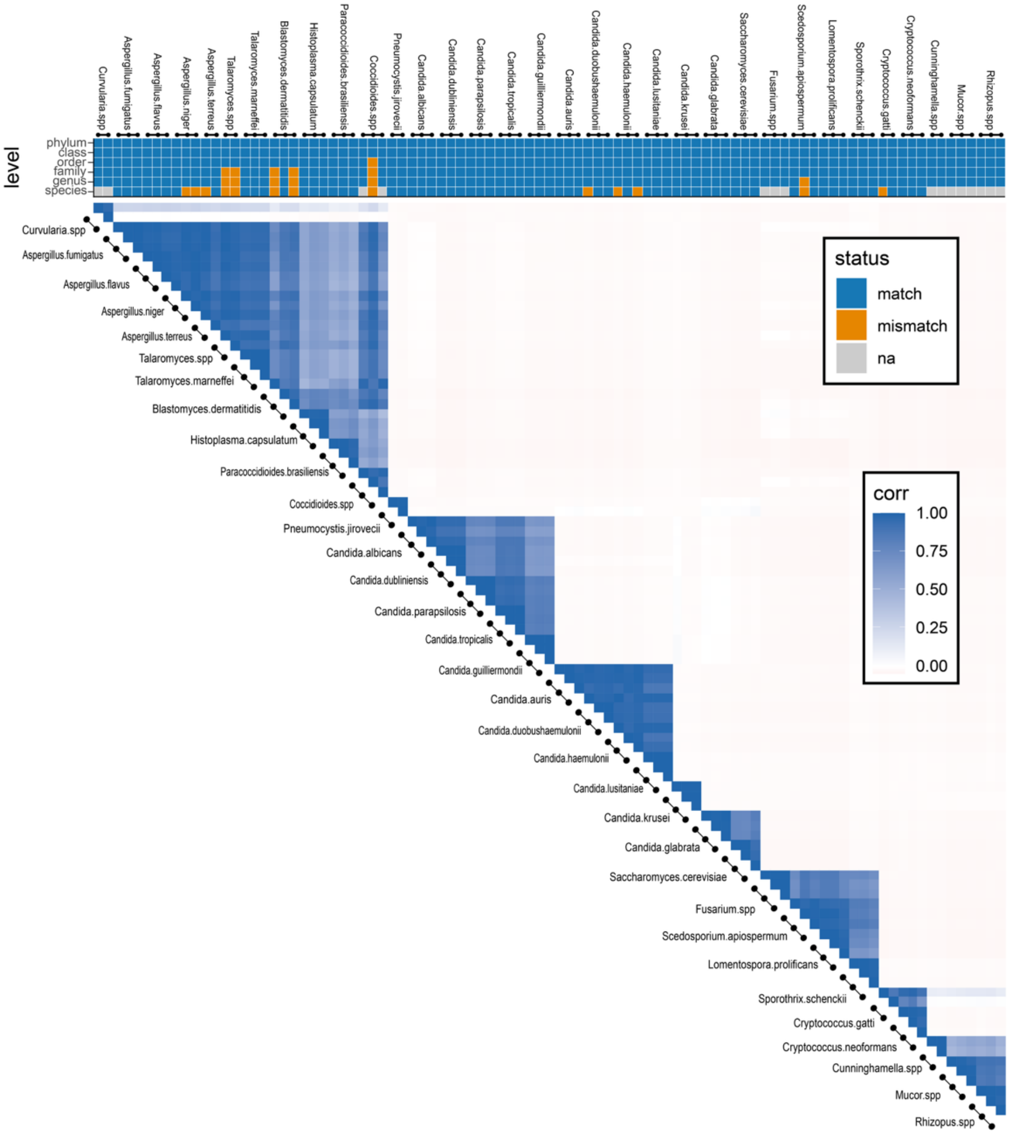


**Figure S3. Pearson correlations of Training Set PSRPs are highest between closely related species.** Samples with similar taxonomic classification had higher pairwise Pearson correlations than those that were more dissimilar. **(A)** Tile plot indicates prediction accuracy for the single best-matched (highest) non-self-Pearson correlation at each taxonomic level, as in Fig 3a. **(B)** Heatmap of pairwise Pearson correlations, showing that the highest correlations map to members of closely related species. Training Set samples are ordered by taxonomic classification as in Fig 2.

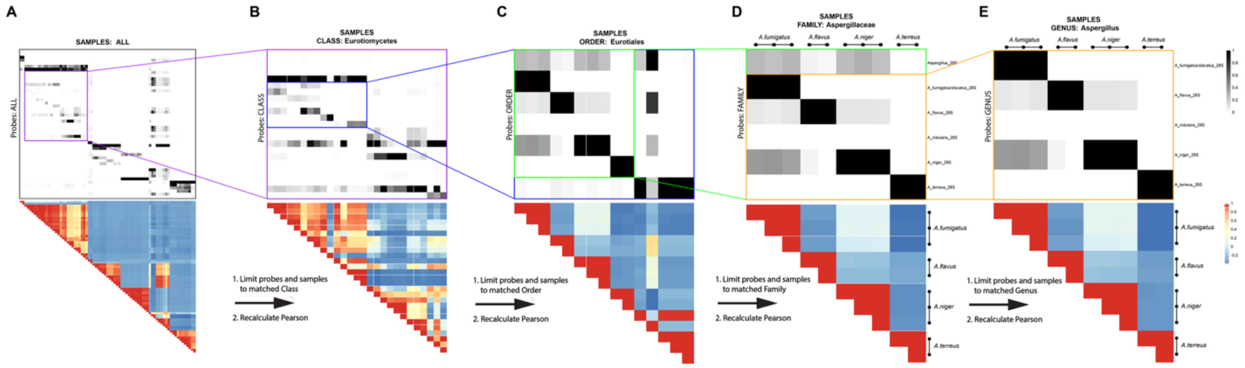


**Figure S4. Schematic of our hierarchical Pearson correlation classifier Co-PILOT.** Visual representation of how Co-PILOT moves through taxonomic levels, demonstrated on an *Aspergillus fumigatus* sample. Co-PILOT begins by considering all probes and samples **(A)** and chooses the best-matched class based on the class of the single (non-self) sample with the highest Pearson correlation to the query sample. It then restricts further analysis to only samples from that class, and only probes nested within that class (i.e., those designed to target regions matching an order, family, genus, or species within the selected class), indicated by the box (colored by taxonomic level, as in other figures; here, purple box = class); data from all other samples and probes are removed at this stage **(B)**, including probe(s) matching selected class. In subsequent steps, Co-PILOT selects only the subset of samples & probe(s) containing the best-matched order (**B** → **C**, blue box), family (**C** → **D**, green box), and genus (**D** → **E**, orange box). Top panels = read count heatmaps, rescaled by sample at each step to maximize remaining probe; bottom panels = Pearson correlation heatmaps only using selected data at each step.

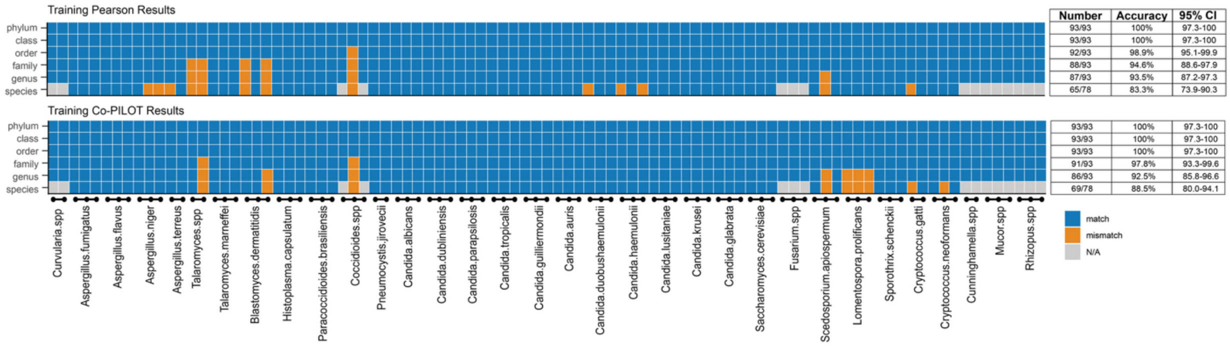


**Figure S5. The Co-PILOT classifier improved identification accuracy of the Pan-Fungal Phirst-ID probeset on the Training Set.** Tile plot indicating prediction accuracy at each taxonomic level, as in Fig 3a. Co-PILOT (bottom panel) improved accuracy relative to simple Pearson correlations (top panel), though it was iteratively designed based on the Training Set, so these results are best viewed as “overtrained” - see main text and Methods for details. By contrast, its performance on the Validation Set (Fig 6) is an independent assessment of Co-PILOT’s accuracy on data it had not seen until its algorithm (Supp Fig S4) was fully designed.

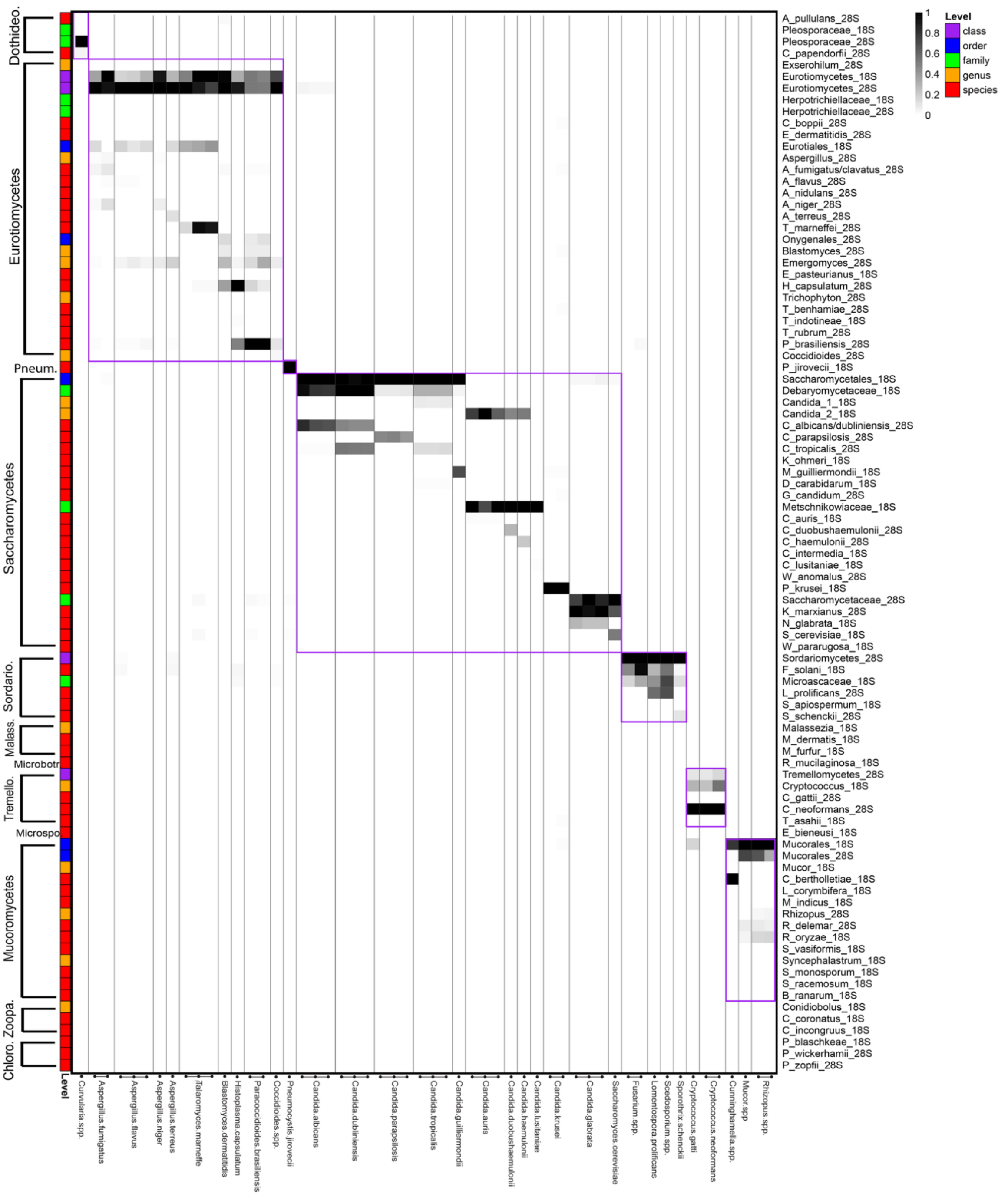


**Figure S6. Pan-Fungal Phirst-ID panel generates unique PSRPs for 32 clinically relevant species in our Validation Set.** Heatmap of scaled binding intensities of the 54 samples comprising our Validation Set. As in Figure 2, samples and probes are ordered taxonomically, though the cladogram is omitted here. Taxonomic level of each probe is indicated by colored box at left; probe name is listed at right. Purple boxes indicate regions where the class of samples and intended probe targets match.

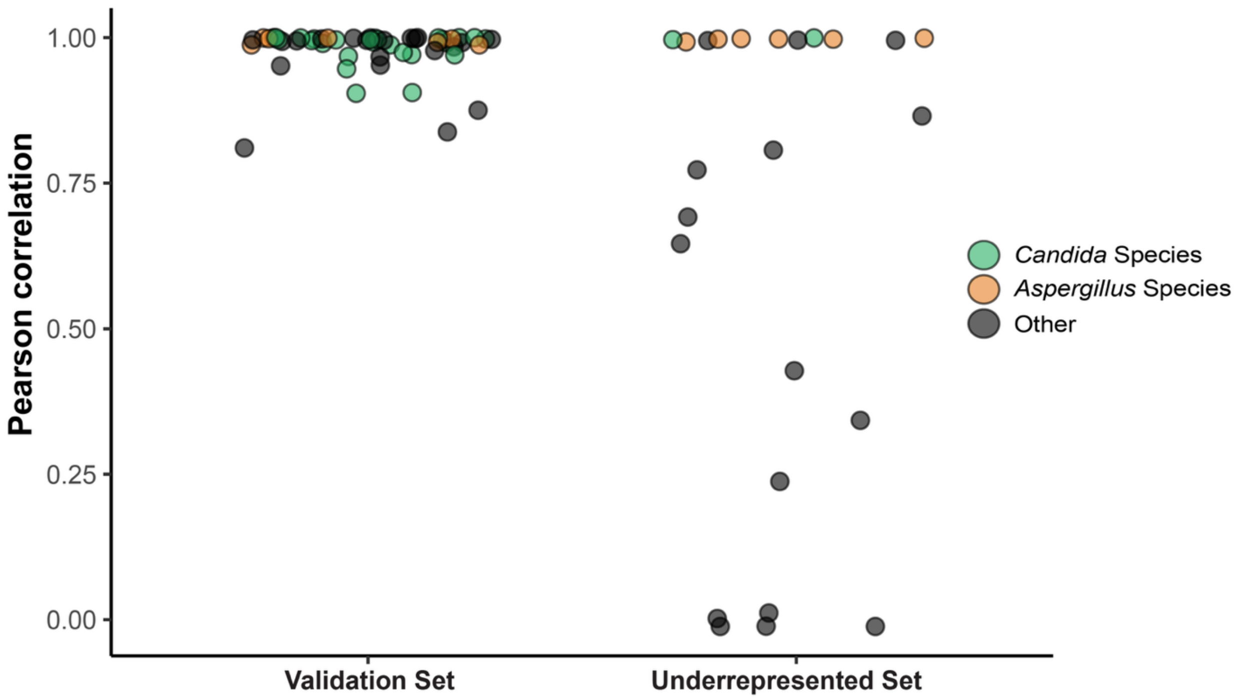


**Figure S7. Pearson correlations of best match to Training Set for previously seen and unseen species.** Dotplot showing Pearson correlations of test samples against the Training Set reference profile. Samples are grouped by whether their species is represented in the Training Set. The Validation Set (left) contains samples present in the Training Set, whereas the Underrepresented Set (right) contains samples with no matching species. Point color denotes taxonomic groups highlighting *Candida* and *Aspergillus* species in each Set.

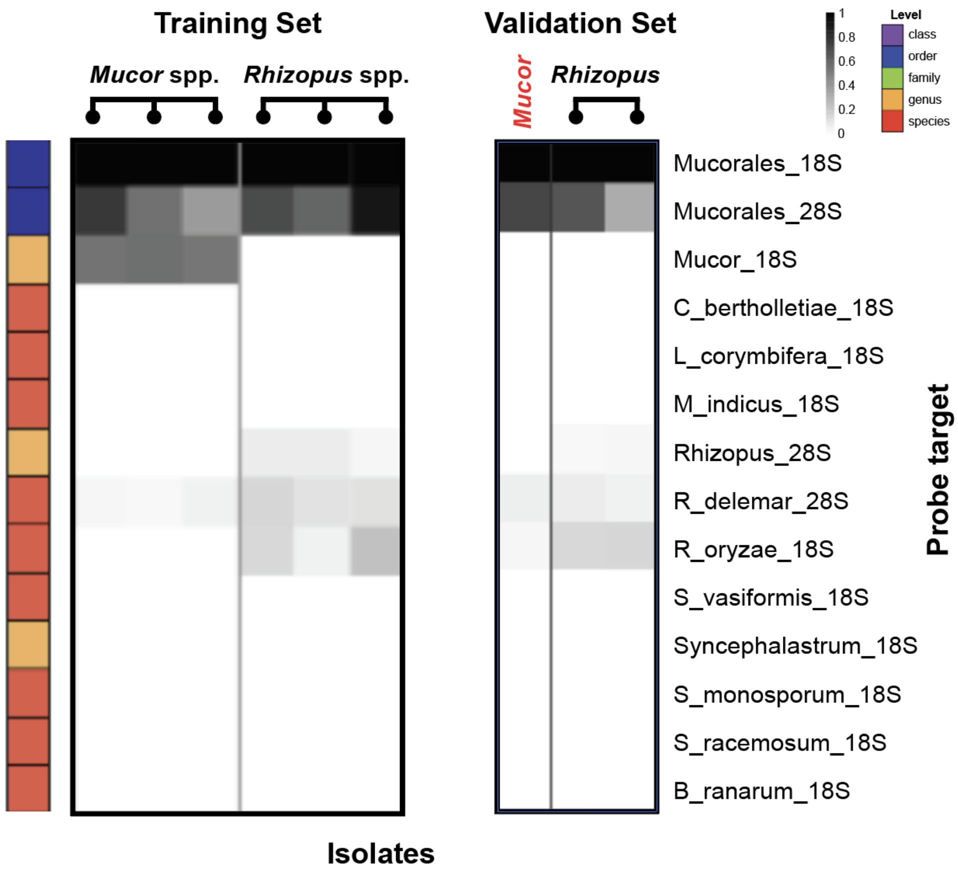


**Figure S8. Errant clinical identification of *Mucor* from Validation Set.** Pan-Fungal Phirst-ID data from all probes targeting subsets of the *Mucoromycetes* class, shown for all samples from the Training and Validation Sets identified by the clinical microbiology laboratory as either *Mucor* or *Rhizopus*. The red label indicates a Validation sample identified by the clinical laboratory as a *Mucor*, but with a PSRP better resembling a *Rhizopus* (specifically in the lack of Mucor_18S probe reactivity, and perhaps greater R_oryzae_18S probe reactivity). ITS sequencing identified this sample as *Rhizopus microsporus*, more consistent with the Pan-Fungal Phirst-ID prediction than the clinical classification.

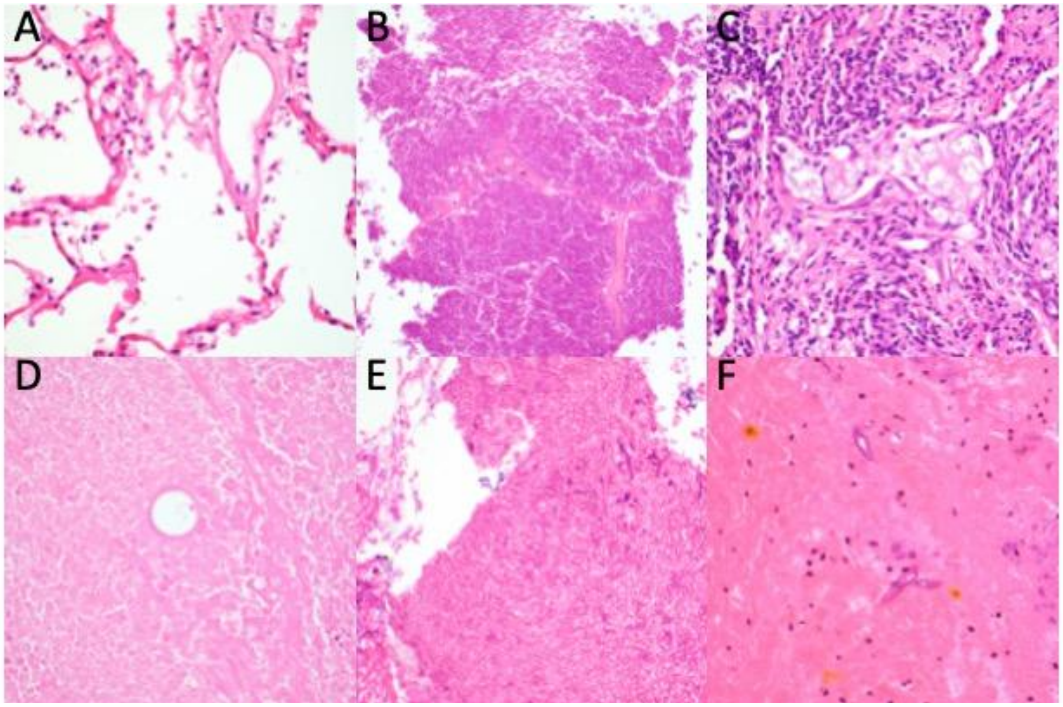


**Figure S9. Representative hematoxylin and eosin (H&E)-stained sections from formalin-fixed, paraffin-embedded tissues included in the study. (A)** Lung tissue, negative control (Fig S9, right panel, column 11; Supp Table S3d, sample FP002). **(B)** Aortic valve tissue with *Candida parapsilosis* (Fig S9, left panel, column 7; Supp Table S3d, sample FP024. **(C)** Lung tissue with *Cryptococcus neoformans* (Fig S9, right panel, column 8; Supp Table S3d, sample FP011). **(D)** Lung tissue with *Coccidioides* sp (Fig S9, right panel, column 4; Supp Table S3d, sample FP019. **(E)** Sinus tissue with *Aspergillus fumigatus* (Fig S9, left panel, column 5; Supp Table S3d, sample FP028). **(F)** Sinus tissue with *Rhizopus* sp (Fig S9, right panel, column 15; Supp Table S3d, sample FP023). All images were acquired using a 40× objective.

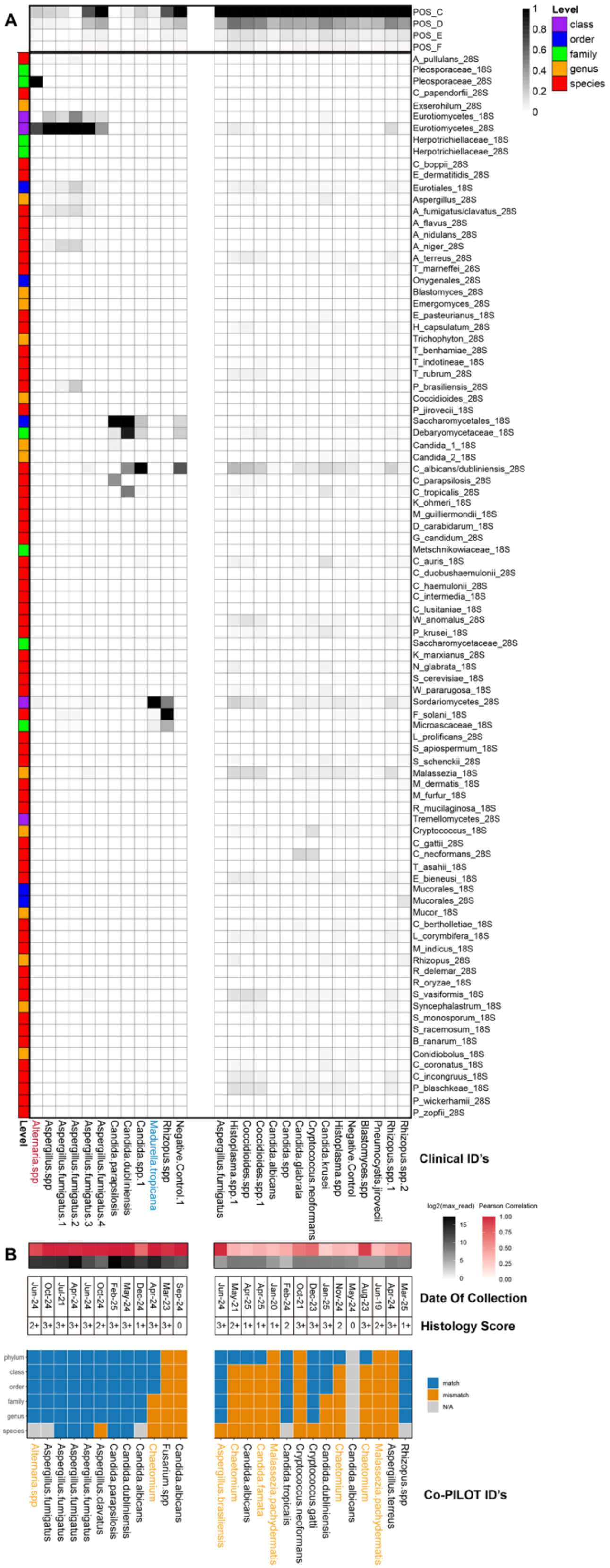
A
B
POS_C
POS_D
POS_E
POS_F
A_pullulans_28S
Pleosporaceae_18S
Pleosporaceae_28S
C_papendorfii_28S
Exserohilum_28S
Eurotiomycetes_18S
Eurotiomycetes_28S
Herpotrichiellaceae_18S
Herpotrichiellaceae_28S
C_boppii_28S
E_dermatitidis_28S
Eurotiales_18S
Aspergillus_28S
A_fumigatus/clavatus_28S
A_flavus_28S
A_nidulans_28S
A_niger_28S
A_terreus_28S
T_marneffei_28S
Onygenales_28S
Blastomyces_28S
Emergomyces_28S
E_pasteurianus_18S
H_capsulatum_28S
Trichophyton_28S
T_benhamiae_28S
T_indotineae_18S
T_rubrum_28S
P_brasiliensis_28S
Coccidioides_28S
P_jirovecii_18S
Saccharomycetales_18S
Debaryomycetaceae_18S
Candida_1_18S
Candida_2_18S
C_albicans/dubliniensis_28S
C_parapsilosis_28S
C_tropicalis_28S
K_ohmeri_18S
M_guilliermondii_18S
D_carabidarum_18S
G_candidum_28S
Metschnikowiaceae_18S
C_auris_18S
C_duobushaemulonii_28S
C_haemulonii_28S
C_intermedia_18S
C_lusitaniae_18S
W_anomalus_28S
P_krusei_18S
Saccharomycetaceae_28S
K_marxianus_28S
N_glabrata_18S
S_cerevisiae_18S
W_pararugosa_18S
Sordariomycetes_28S
F_solani_18S
Microascaceae_18S
L_prolificans_28S
S_apiospermum_18S
S_schenckii_28S
Malassezia_18S
M_dermatis_18S
M_furfur_18S
R_mucilaginosa_18S
Tremellomycetes_28S
Cryptococcus_18S
C_gattii_28S
C_neoformans_28S
T_asahii_18S
E_bieneusi_18S
Mucorales_18S
Mucorales_28S
Mucor_18S
C_bertholletiae_18S
L_corymbifera_18S
M_indicus_18S
Rhizopus_28S
R_delemar_28S
R_oryzae_18S
S_vasiformis_18S
Syncephalastrum_18S
S_monosporum_18S
S_racemosum_18S
B_ranarum_18S
Conidiobolus_18S
C_coronatus_18S
C_incongruus_18S
P_blaschkeae_18S
P_wickerhamii_28S
P_zopfii_28S
1
0.8
0.6
0.4
0.2
0
Level
class
order
family
genus
species
Level
Alternaria.spp
Aspergillus.spp
Aspergillus.fumigatus.1
Aspergillus.fumigatus.2
Aspergillus.fumigatus.3
Aspergillus.fumigatus.4
Candida.parapsilosis
Candida.dubliniensis
Candida.spp.1
Madurella.tropicana
Rhizopus.spp
Negative.Control.1
Aspergillus.fumigatus
Histoplasma.spp.1
Coccidioides.spp
Coccidioides.spp.1
Candida.albicans
Candida.spp
Candida.glabrata
Cryptococcus.neoformans
Candida.krusei
Histoplasma.spp
Negative.Control
Blastomyces.spp
Pneumocystis.jiroveci
Rhizopus.spp.1
Rhizopus.spp.2
Clinical ID's
log2(max_read)
15
10
5
0
Pearson Correlation
1.00
0.75
0.50
0.25
0.00
Jun-24
Oct-24
Jul-21
Apr-24
Jun-24
Oct-24
Feb-25
May-24
Dec-24
Apr-24
Mar-23
Sep-24
Jun-24
May-21
Apr-25
Apr-25
Jan-20
Feb-24
Oct-21
Dec-23
Jan-25
Nov-24
May-24
Aug-23
Jun-19
Apr-24
Mar-25
Date Of Collection
2+
3+
3+
3+
3+
2+
3+
3+
1+
3+
3+
0
3+
2+
1+
1+
1+
2
3+
3+
3+
2
0
3+
2+
3+
1+
Histology Score
phylum
class
order
family
genus
species
match
mismatch
N/A
Alternaria.spp
Aspergillus.fumigatus
Aspergillus.fumigatus
Aspergillus.fumigatus
Aspergillus.fumigatus
Aspergillus.clavatus
Candida.parapsilosis
Candida.dubliniensis
Candida.albicans
Chaetomium
Fusarium.spp
Candida.albicans
Aspergillus.brasiliensis
Chaetomium
Candida.albicans
Candida.famata
Malassezia.pachydermatis
Candida.tropicalis
Cryptococcus.neoformans
Cryptococcus.gattii
Candida.dubliniensis
Chaetomium
Candida.albicans
Chaetomium
Malassezia.pachydermatis
Aspergillus.terreus
Rhizopus.spp
Co-PILOT ID's

**Figure S10: Pilot use of Pan-Fungal Phirst-ID for FFPE samples. (A)** Heatmap of scaled probe binding intensities of 27 FFPE samples. For column-scaling to better visually represent total read counts, the four lowest-abundance positive control spike-ins are included for each sample (top 4 rows, above horizontal line). Bottom axis indicates clinical identification provided by the Pathology department (see Supplementary Table S3d). Red text indicates an organism not included in probeset design, blue text indicates an organism neither included in probeset design for nor previously tested by Pan-Fungal Phirst-ID (and thus not available to the Co-PILOT algorithm for comparison). **(B)** Tile plot indicating prediction accuracy of Pan-Fungal Phirst-ID with Co-PILOT at each taxonomic level, as in Fig 3a. Date of sample collected is indicated above and final Co-PILOT identification (best match from Training Set) is listed below the tile plot. 12 samples at left, which exceeded our probe detection threshold, are separated from 17 samples at right, which did not. Samples are sorted by taxonomic order of the identification from the clinical microbiology laboratory. Orange text indicates a Co-PILOT prediction of a species not included in probeset design.
